# *IFT74* variants cause skeletal ciliopathy and motile cilia defects in mice and humans

**DOI:** 10.1101/2023.02.23.23286106

**Authors:** Zeineb Bakey, Oscar A. Cabrera, Julia Hoefele, Dinu Antony, Kaman Wu, Michael W. Stuck, Dimitra Micha, Thibaut Eguether, Abigail O. Smith, Nicole N. van der Wel, Matias Wagner, Lara Strittmatter, Philip L. Beales, Julie A. Jonassen, Isabelle Thiffault, Maxime Cadieux-Dion, Laura Boyes, Saba Sharif, Beyhan Tüysüz, Desiree Dunstheimer, Hans W.M. Niessen, William Devine, Cecilia W Lo, Hannah M. Mitchison, Miriam Schmidts, Gregory J. Pazour

**Affiliations:** Center for Pediatrics and Adolescent Medicine, University Hospital Freiburg, Freiburg University Faculty of Medicine, Mathildenstrasse 1, 79112 Freiburg, Germany; Human Genetics Department, Radboud University Medical Center Nijmegen and Radboud Institute for Molecular Life Sciences (RIMLS) Nijmegen, Geert Grooteplein Zuid 10, 6525KL, The Netherlands; Program in Molecular Medicine, University of Massachusetts Chan Medical School, Biotech II, Suite 213, 373 Plantation Street, Worcester, MA 01605, USA; Institute for Human Genetics, Technical University Munich (TUM), School of Medicine, Trogerstraße 32 81675 Munich, Germany; Department of Human Genetics, Amsterdam Movement Sciences, Amsterdam UMC, De Boelelaan 1117, 1081 HV Amsterdam, The Netherlands; Electron microscopy Center Amsterdam, Department of Medical Biology, VUMC, 1105 AZ, Amsterdam, The Netherlands; Electron Microscopy Core, University of Massachusetts Chan Medical School, 55 Lake Avenue North, Worcester, MA 01655, USA; Genetics and Genomic Medicine Programme, University College London, UCL Great Ormond Street Institute of Child Health, London WC1N 1EH, UK; Department of Microbiology and Physiological Systems, University of Massachusetts Chan Medical School, 55 Lake Avenue North, Worcester MA 01655, USA; Genomic Medicine Center, Children’s Mercy Hospital, 2401 Gillham Road, Kansas City, MO 64108, USA; West Midlands Genomic Medicine Hub, Birmingham Women’s Hospital, Mindelsohn Way, Birmingham B15 2TG, UK; Department of Pediatrics, Division of Pediatric Genetics, Cerrahpasa Medical Faculty, University-Cerrahpasa, 34303 Istanbul, Turkey; Center for Pediatrics and Adolescent Medicine, University Hospital Augsburg, 86156 Augsburg, Germany; Department of Pathology, Amsterdam University Medical Center (AUMC), Amsterdam, The Netherlands; Department of Developmental Biology, University of Pittsburgh, 8111 Rangos Research Center, 530 45th Street Pittsburgh, PA 15201, USA; CIBSS—Center for Integrative Biological Signaling Studies, University of Freiburg, 79104 Freiburg, Germany

## Abstract

Motile and non-motile cilia are critical to mammalian development and health. Assembly of these organelles depends on proteins synthesized in the cell body and transported into the cilium by intraflagellar transport (IFT). A series of human and mouse *IFT74* variants were studied to understand the function of this IFT subunit. Humans missing exon 2, which codes for the first 40 residues, presented an unusual combination of ciliary chondrodysplasia and mucociliary clearance disorders while individuals carrying biallelic splice site variants developed a lethal skeletal chondrodysplasia. In mice, variants thought to remove all Ift74 function, completely block ciliary assembly and result in midgestational lethality. A mouse allele that removes the first 40 amino acids, analogous to the human exon 2 deletion, results in a motile cilia phenotype with mild skeletal abnormalities. *In vitro* studies suggest that the first 40 amino acids of IFT74 are dispensable for binding of other IFT subunits but are important for tubulin binding. Higher demands on tubulin transport in motile cilia compared to primary cilia could account for the motile cilia phenotype observed in human and mice.

## Introduction

Cilia are evolutionarily conserved, microtubule-based organelles that serve diverse sensory and motility functions throughout the eukaryotic kingdom. In mammals and other vertebrates, most cell types display non motile primary cilia. These cilia are thought to sense the extracellular milieu allowing cells to coordinate with their environment, which is critical during development and adult tissue homeostasis (1, 2). In addition, certain specialized cells display motile cilia that can move fluids over their surface and or power their movement though liquids. These cilia play critical roles in maintaining lung health and male fertility, and also are critical in the establishment of the left-right pattern of the heart and other organs.

With the diversity of ciliary structure and function though out the body, it follows that disease-causing variants (likely pathogenic and pathogenic variants) in different ciliary genes can cause strikingly different phenotypes. Typically, dysfunction of primary cilia causes complex developmental phenotypes affecting multiple organs. The organ involvement varies but commonly renal, liver and pancreatic cysts are observed along with retinal degeneration, brain abnormalities and skeletal malformations including polydactyly, brachydactyly, shortened long bones and short ribs (3, 4). In humans and mice, motile cilia dysfunction results in male infertility, hydrocephaly, primary ciliary dyskinesia (PCD) / mucociliary clearance disorders and disturbances of the left-right body axis. Male infertility is thought to result from impaired sperm motility while hydrocephaly is thought to result from impaired movement of cerebrospinal fluid over the ependymal cells of the brain. PCD is characterized by reduced mucociliary clearance of the airways causing recurrent respiratory tract infections, ultimately leading to irreversible pulmonary dysfunction. Disrupted left-right patterning can result in internal organ reversal (situs inversus) or randomization (heterotaxy) (5-7). Situs inversus is a relatively benign condition whereas heterotaxy often causes complex structural heart disease (5, 8-14).

As in most eukaryotes, mammalian ciliary assembly is driven by intraflagellar transport (IFT) (15, 16). During IFT, kinesin-2 and dynein-2 motors transport large multimeric IFT trains from the cell body into and along the ciliary microtubules. The IFT trains are composed of three evolutionarily conserved subcomplexes, IFT-A, IFT-B and the BBSome. IFT-A contains 6 proteins (IFT144, IFT140, IFT121, IFT120, and IFT43), IFT-B contains 16 proteins (IFT172, IFT88, IFT81, IFT80, IFT74, IFT70, IFT56, IFT54, IFT57, IFT52, IFT46, IFT38, IFT27, IFT25, IFT22, IFT20) and the BBSome contains eight proteins (BBS1, BBS2, BBS4, BBS5, BBS7, BBS8, BBS9 and BBIP1). While much remains to be understood about cargo binding, studies have indicated that IFT-A working with Tulp3 is important for the delivery of membrane proteins to cilia (17), IFT46 working with ODA16 delivers outer dynein arm components (18, 19) and IFT74/IFT81 deliver tubulin to the organelle (20, 21).

Disease-causing variants in at least five IFT-A, six IFT-B and five dynein-2 genes cause congenital syndromes with skeletal, renal, and retinal defects (22, 23). These are clinically grouped as ciliary chondrodysplasias but vary in severity. Short-rib polydactyly (SRPS) is at the severe end of the spectrum and is incompatible with survival beyond the neonatal period (24). Jeune asphyxiating thoracic dystrophy (ATD) presentation is variable ranging from cardiorespiratory lethality due to lung hypoplasia in ∼20% of cases to milder skeletal phenotypes depending on the underlying genetic cause. Interestingly, hypomorphic IFT-gene dysfunction in ATD or Cranioectodermal dysplasia (CED) causes frequent extraskeletal renal, hepatic and retinal disease in contrast to hypomorphic dynein-2 complex variants causing a severe phenotype restricted to the skeleton (25). The pulmonary hypoplasia is thought to be a consequence of lung growth restriction *in utero* caused by the abnormally small thoracic cavity due to the shortened ribs; however, lung-patterning defects may contribute. Respiratory distress is shared with mucociliary clearance disorders, including Primary Ciliary Dyskinesia (PCD) but mucociliary clearance disorders are not usually associated with skeletal, kidney or retinal defects. Respiratory failure caused by motile ciliopathies develops slowly over several decades due to recurrent respiratory infections. Currently more than 60 mucociliary clearance disorder disease genes have been identified. These genes encode subunits of the motility apparatus, their assembly factors, or genes involved in multiciliogenesis but in general are not needed for assembly of non-motile primary cilia (26, 27).

Here we report human genetic variants in the gene encoding the IFT74 subunit of IFT-B. In four independent families, the variant in *IFT74* caused an ATD/SRPS-like skeletal dysplasia spectrum phenotype. Two of these families carry a novel deletion of exon 2 and present with PCD phenotypes in addition to ATD-like skeletal dysplasia features. The other two families carry splicing alleles associated with a SRPS phenotype with congenital heart defects. Mouse models showed that a complete loss of *Ift74* prevented ciliary assembly and caused severe cardiac malformations with complete lethality at about embryonic day 9. A mouse allele similar to the human exon 2 deletion formed primary cilia but had defects in the assembly of motile cilia. These mice survived gestation and were born alive but showed growth restriction in the postnatal period and developed hydrocephaly.

## Results

### Identification of individuals carrying biallelic *IFT74* loss of function variants

To identify genetic causes of ATD, SRPS and ATD-like ciliary chondrodysplasia phenotypes, we performed exome and targeted capture sequencing of known skeletal ciliopathy and PCD genes from affected individuals. All individuals or their legal representatives have consented to take part in this study and consent for publication of images has been obtained where applicable. In four individuals from four unrelated families, we found biallelic putative loss of function variants in *IFT74*. Sanger sequencing confirmed a biallelic loss of function variant in a clinically affected sibling in one of the families (Figure 1, S1; Table 1, S1).

**Figure 1.**
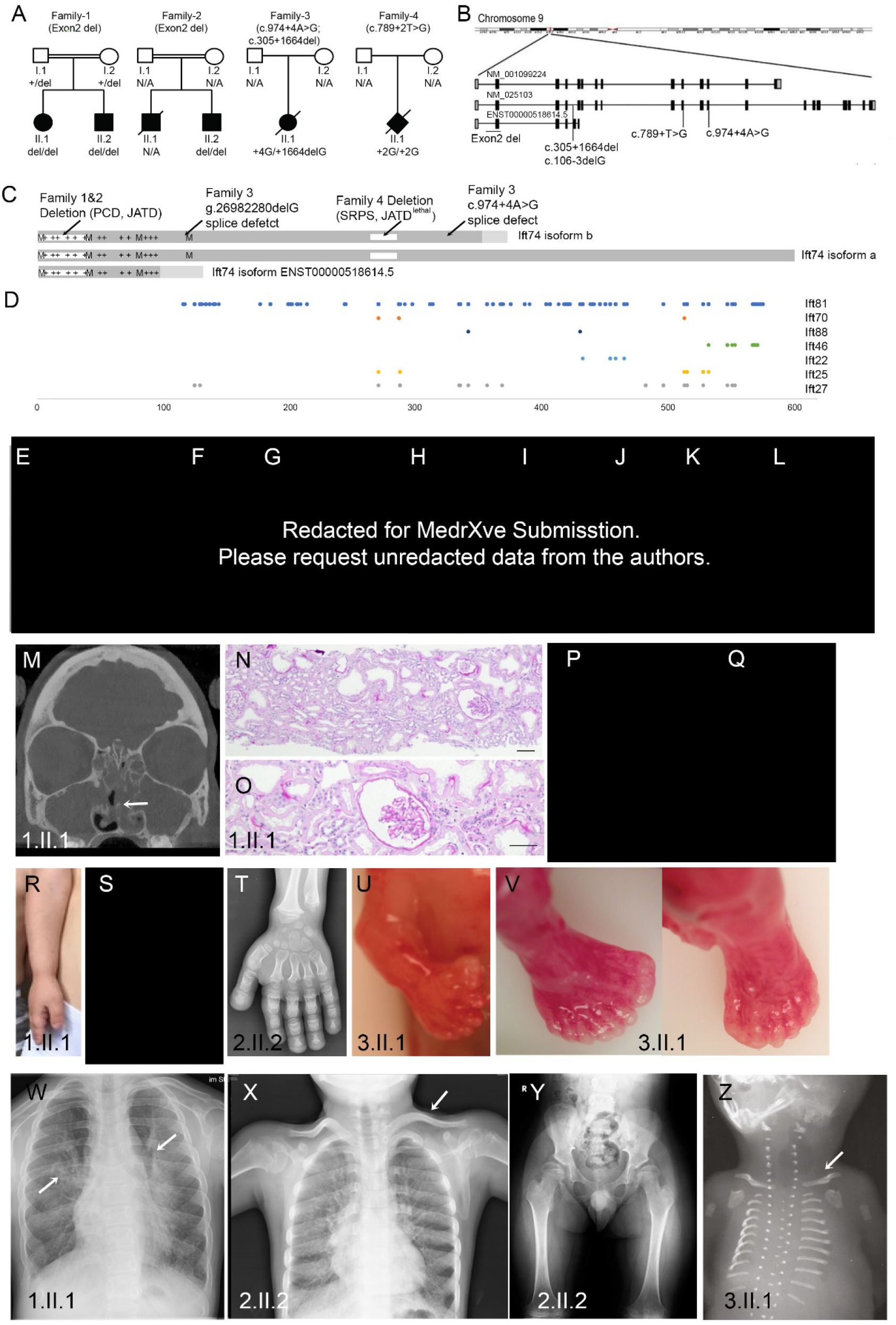
IFT74 Patients Develop Ciliopathy Phenotypes. **A**. Family trees of affected individuals. **B**. Diagram of the IFT74 locus showing the positions of NM_025103, NM_001099224, and ENST00000518614.5 transcripts. **C**. IFT74 protein structure. Isoform a is composed of 600 residues, isoform b 372 residues and ENST00000518614.5 146 residues. The N-terminal 351 residues of isoform b are identical to isoform a, and there are 21 unique residues at the C-terminal end. ENST00000518614.5 shares 102 residues with isoforms a and b, and has 44 unique residues at its C-terminal end. The basic, tubulin binding domain at the N-terminus, is marked with +’s to show positions of lysines and arginines. The first four methionines (M) at positions 1,40, 80, and 121 are shown (mouse Ift74 also has a methionine at position 84). The locations of the deletions of 1-40 in family 1and 2, and 263-274 in family 4 are shown. **D**. Crosslinks to IFT74 by other IFT-B proteins are shown at the bottom. These were identified in *Chlamydomonas* IFT74 (39) and mapped to corresponding positions in human IFT74 isoform a. **E, G, K**. Narrow thorax and shortened upper extremities in all three affected individuals, most pronounced in 1.II.1. **F-K**. Shortened lower extremities in all three individuals with pronounced bowing of the lower legs. **L**. Severely shorted extremities and narrow thorax in fetus 3.II.1. **M**. Head CT image of patient 1.II.1 showing a fluid filled/mucus blocked sinus (arrow). **N-O**. Hematoxylin and eosin-stained kidney biopsy from 1.II.1 showing mild tubular dilatations and a small glomerular cyst. Scale bars are 100 microns. **P-Q**. Growth curves of patients 1.II.1 and 1.II.2 showing progressive growth retardation with age. **R-T**. Severe brachydactyly with shortened metacarpal bones. **U**. Pre-axial polydactyly of the right hand **V**. Pre- and postaxial polydactyly of the right foot and postaxial polydactyly of the left foot. **W**. Chest radiograph showing pulmonary infiltrations (arrows). **X**. Chest x-ray showing handlebar clavicles (arrow) and shortened horizontal ribs. **Y**. Pelvis x-ray showing a narrow pelvis with a coxa valga deformity. **Z**. Chest x-ray showing very short horizontal ribs and handlebar clavicles (arrow).

**Table 1:** Clinical Data.

| Feature | Family 1 |  | Family 2 | Family 3 | Family 4 |
| --- | --- | --- | --- | --- | --- |
| Case | 1.II.1 | 1.II.2 | 2.II.2 | 3.II.1 | 4.II.1 |
| Current age | Adolescent (female) | Child (male) | Adolescent (male) | Fetus (female) | Neonate, (male) deceased |
| Clinical diagnosis | Jeune asphyxiating thoracic dystrophy / mucociliary clearance disorder | Jeune asphyxiating thoracic dystrophy / mucociliary clearance disorder | Jeune asphyxiating thoracic dystrophy / mucociliary clearance disorder | ATD / SRPS | SRPS/Jeune asphyxiating thoracic dystrophy |
| Genetic analysis performed | ES with CNV analysis followed by GS | Sanger sequencing | ES with CNV analysis | NGS panel | ES |
| *IFT74* variants (NM_025103.4/ENST00000380062.9, GRCh38/hg38) | Homozygous exon 2 deletion | Homozygous exon 2 deletion | Homozygous exon 2 deletion | Heterozygous splice donor variant, c.974+4A>G (intron 12) + c.305+1664delG (intron 4) | Homozygous splice donor variant c.789+2T>G (intron 10) |
| Short ribs/narrow chest | Yes | Yes | Yes | Yes | Restrictive pulmonary hypoplasia |
| Polydactyly | No | No | No | Yes, multiple polysyndactyly affecting all (both hands, both feet) | Affecting all 4 extremities |
| Disproportional short stature and skeletal features | -5.9 SD<br>More pronounced with age, bowing of legs. Achondroplasia-like, short limbs, brachydactyly, | -4.9 SD<br>Less severe than I.II.1; more pronounced with age, short limbs, brachydactyly | -8.5 SD (52 cm at birth, final height 118 cm)<br>Short stature more pronounced with age, narrow thorax, short limbs, brachydactyly. Mild pectus excavatus, slanted acetabulum, hypermobility | Short proximal and distal limbs, severely short femurs/humeri. Short irregular growth plates, broad, poorly ossified cartilage-rich trabeculae, small scapulae and iliacs. 11 rib pairs, poorly ossified long, hand and foot bones and vertebral bodies. | Severely shortened extremities, thoracic narrowing. |
| Cranial and facial defects | Narrow foramen magnum | No | Flat nose, high arched palate. | Low set ears, broad nose, micrognathia. Midline cleft of upper lip. Posterior midline cleft palate, lobulated anterior tongue, tongue tie. Large head with wide skull sutures, absent brain falx and corpus callosum. | No |
| Heart defect | Mitral valve insufficiency, ASD II, PDA | No | Double aortic arch. (Older sib died of heart defect) | Persistent left superior vena cava, AVSD, hypoplastic left heart, aortic atresia. | DORV, hypoplastic left ventricle |
| Renal disease | Progressive renal insufficiency. dialysis started as a teenager | No | Hyperechoic kidneys, proteinuria chronic renal insufficiency, arterial hypertension | Cloacal abnormality with fusion of vaginal and bladder and bladder outflow obstruction. Dilated ureters and renal pelvis; bilateral renal cystic dysplasia on histology. | No. |
| Liver Disease | No | No | No | Liver histology suggests probable ductal plate malformation. | No |
| Retinal disease | No | No | Retinitis pigmentosa | N/A (fetus) | Not tested |
| Obesity | No | No | Yes | N/A (fetus) | No |
| Intellectual delay | No | No | No | N/A (fetus) | N/A (died after birth as a preterm neonate). |
| Other features | No | No | Hypertension | Annular pancreas | No |
| Situs inversus | No | No | No | Intestinal malrotation. | No |
| Low nNO<br>(normal >300nl/min;<br>in PCD often ><br>77nl/min) | < 150 nl/min | < 150 nl/min | Not tested | N/A (fetus) | Not tested |
| Recurrent<br>respiratory<br>infections | Yes, more severe than<br>I.II.2 | Yes | Recurrent bacterial<br>infections and<br>pneumonia in<br>childhood | N/A (fetus) | N/A |
| Other respiratory<br>features | Neonatal respiratory<br>insufficiency in I.II.1;<br>Recurrent rhinosinusitis<br>and persistent wet cough,<br>more severe in I.II.1 than<br>I.II.2 | Recurrent rhinosinusitis<br>and persistent wet cough | Rhinosinusitis | N/A (fetus) | Severe<br>respiratory<br>insufficiency due<br>to pulmonary<br>hypoplasia |
| Otitis media | Yes, more severe in<br>I.II.1 | Yes | Yes | N/A (fetus) | N/A (died after<br>birth as a<br>preterm<br>neonate). |
| Motile cilia function | Reduced cilia numbers,<br>very short cilia | Reduced cilia numbers,<br>very short cilia, also after<br>cell culturing | Reduced cilia<br>numbers, very short<br>cilia | N/A (fetus) | Not tested. |

NGS, next-generation sequencing; ES, exome sequencing; GS, genome sequencing; SD, standard deviation of
height; ASD, atrial septal defect; PDA, patent ductus arteriosus; AVSD, atrioventricular septal defect; DORV,
double outlet right ventricle; nNO, nasal nitric oxide levels; N/A, not available.

In Family 1 and 2, routine filtering of the exome sequence (28-31) did not show any variants of interest. However, copy number variant analysis of next-generation sequencing data using ExomeDepth software (32) showed that the affected children carried a homozygous deletion of *IFT74* exon 2 (Figure S1A,D). This deletion is absent from the Database of Genomic Variants (33) and not present in ∼10,000 exomes collected at Radboudumc Nijmegen. Genome sequencing performed in individual 1.II.1 confirmed a homozygous genomic re-arrangement with deep intronic breakpoints in intron 2 and intron 3, deleting exon 2 (Chr9:g.26959922_26962977delinsTTATTATACTC) (Figure S1B,C). Sanger sequencing confirmed the variant in a homozygous state in the clinically affected brother 1.II.2 while both parents were found to be heterozygous carriers. Uniprot reports two isoforms of *IFT74* that vary at their C-termini (Figure 1B,C). Exon 2 contains the predicted start codon for both isoforms and the patient deletion is predicted to delete the start codon and first 40 amino acids of both isoforms (Figure 1C). The first codon of exon 3 is a methionine giving potential for in-frame translational initiation at this point.

In Family 3, a fetus with a SRPs phenotype was found to have compound heterozygous splice site variants, c.974+4A>G and g.26982280delG within *IFT74*. Both variants are absent from EVS. c.974+4A>G lies within intron 12 and Alamut software predicts the creation of a novel splice site 4 bp after the original splice which would result in a frameshift allele. g.26982280delG affects a deep intronic position in the two major *IFT74* transcripts. Interestingly, it localizes to the -3 splice acceptor position (c.106-3delG) in a rare transcript ENST00000518614.5 that shares exons 1-4 with the major transcripts but has alternative exons 5 and 6 (Figure 1B). The protein coded by transcript ENST00000518614.5 shares a common 102 residue N-terminus with the two major isoforms followed by 44 unique residues before terminating in the alternative exon 6 (Figure 1C). As no RNA could be obtained from the fetus or the parents and the variants affect non-consensus splice site positions, we performed a minigene assay to test the effects of the variants on splicing. In the minigene assay, c.974+4A>G caused exclusion of exon 12 (41 bp) from the cDNA (Figure S2A). In IFT74 protein, this would be predicted to cause a 13 amino acid deletion followed by frameshift and a premature stop codon (c.Gln325?delfs*). Using sequence carrying the g.26982280delG variant resulted in inclusion of sequence corresponding to the alternative exon 5 of ENST00000518614.5, which was not observed when wild type sequence was used in the assay (Figure S2B). Our finding suggests that the g.26982280delG variant would prevent the formation of major isoforms in favor of the short ENST00000518614.5 isoform, which is not likely to support IFT-B particle formation and cilia assembly.

In Family 4, a newborn with SRPS or lethal ATD was found by exome sequencing to be homozygous for the substitution variant c.789+2T>G. This variant affects the exon 10 consensus donor splice site and is predicted abrogate the splice site (Figure 2B), likely resulting in skipping of exon 10 which would result in an in-frame product with a 21 amino acid deletion after amino acid 263. Inclusion of the entire intron would result in an in-frame insertion of over 1500 amino acid, which seems unlikely. A shift of the splice site to a deeper intronic position could also result in an out of frame product. Unfortunately, no RNA was available from the family for verification.

**Figure 2.**
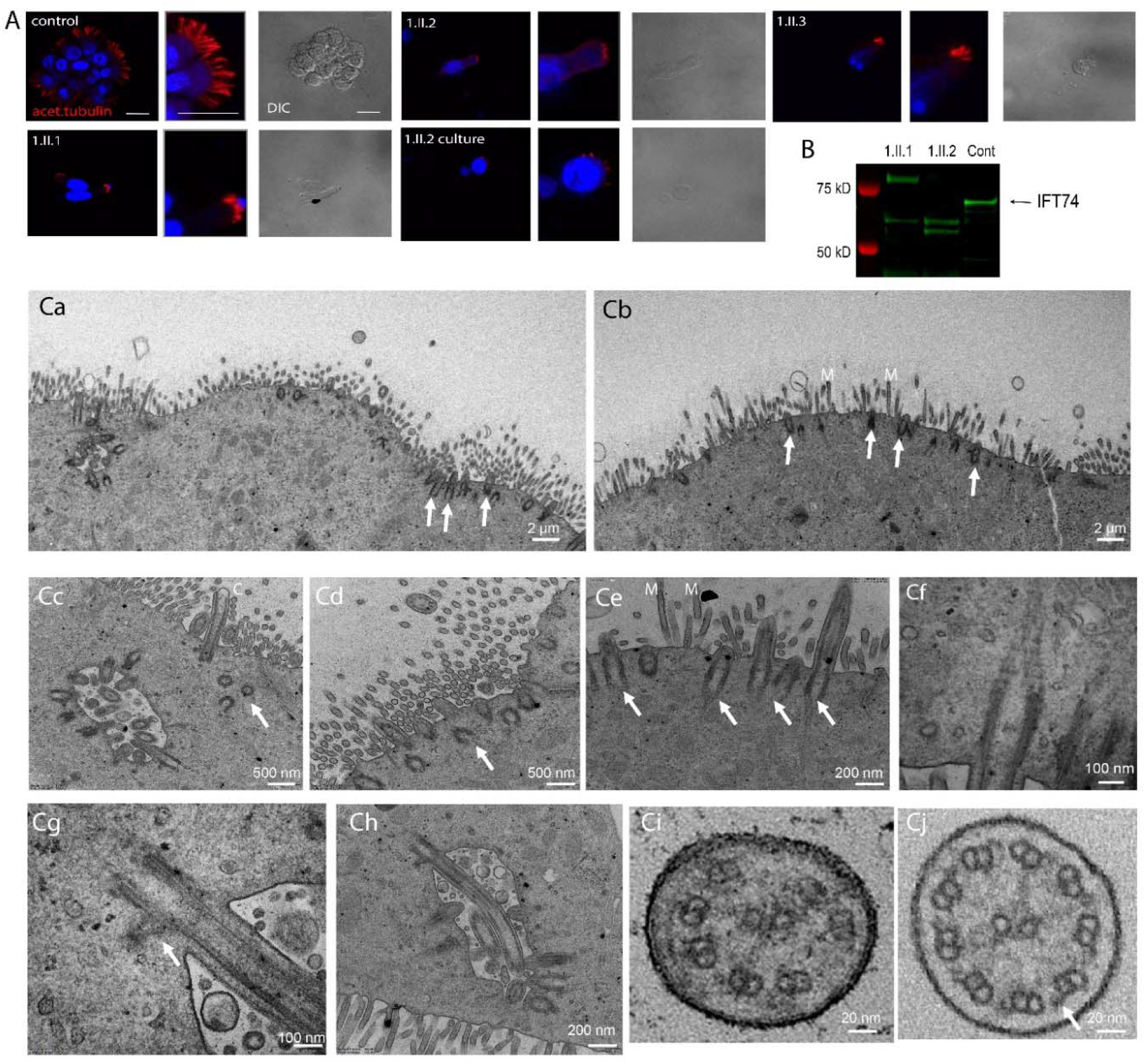
Patients with IFT74exon2Δ Have Motile Cilia Defects. **A**. Immunofluorescence of respiratory epithelium from unaffected patients (control, 1.II.3) and affected (1.II.1 and 1.II.2). 1.II.2 culture was a biopsy that was cultured for 6 weeks before fixation. Red (acetylated tubulin) marks ciliary axonemes. Blue (DAPI) marks nuclei. Scale bars are 20 microns. **B**. Western blot of nasal biopsy samples. **C**. TEM overview images of a multiciliated nasal cells (**Ca**, individual 1.II.2 after cell culture; **Cb**, 1.II.1 native before cell culture) depicting very short cilia (arrows) mostly not even reaching the length of adjacent microvilli (M). **Cc**,**Cd**, close-up image of a ciliated nasal cell in individual 1.II.1 showing shortened cilia (**Cc**) and several undocked basal bodies below the cell surface (arrows). **Ce**, short cilia extending from normal docked basal bodes (arrows) in individual 1.II.1. **Cf**, normal appearing striated ciliary rootlet in 1.II.1. **Cg**, normal appearing striated basal foot in individual 1.II.1. H, example of a longer normally docked cilium in 1.II.1. **Ci**, ciliary cross sections example of individual 1.II.1 showing microtubule pair misarrangement with missing pairs and missing outer dynein arms, indicated with an arrow in a control cross section in **Cj**.

### Phenotypic Analysis of Affected Individuals

Family 1, a consanguineous family, displayed a spectrum of ciliopathy disease features. Individual 1.II.1 presented with a narrow thorax, brachydactyly and short extremities at birth (Figure 1E-G,M-P,R,W). Low thorax volume and recurrent respiratory infections prompted multiple hospital visits with an additional mucociliary clearance disorder clinically suspected. Disproportional short stature of increasing severity with age was noted with a current height at - 5.9 SD (Figure 1P,Q). Severe brachydactyly and development of severe bowing of the lower legs was noted (Figure 1G). The child developed mild proteinuria. Renal biopsy revealed mild tubular and glomerular dilatations with slightly increased interstitial fibrosis (Figure 1N,O). Renal disease progressed during late childhood and end-stage kidney disease with the need for dialysis was reached in adolescence. During childhood, the patient presented with recurrent respiratory infections and nasal nitrous oxide below 200 ppm. Transmission electron microscopic (TEM) analysis of ciliated nasal polyp tissue showed reduced cilia number and cilia length with inconsistent loss of outer dynein arms and translocation of microtubule pairs. Diagnostic genetic analysis for chondrodysplasias, including achondroplasia and targeted next-generation sequencing gene panel analysis for all known ATD, Sensenbrenner Syndrome and PCD genes to date (Bioscentia, Germany) did not reveal any disease-causing variants. Her brother, individual 1.II.2, presented with a milder but similar phenotype of mildly narrowed thorax, short stature worsening with age, brachydactyly and recurrent respiratory infections but no renal disease has been detected (Figure 1G-I,Q).

Individual 2.II.2 was born as second child to a consanguineous family with a congenital heart defect (double aortic arch). The individual displayed a narrow thorax but had normal birth length (52 cm) and developed a progressive short stature and progressive limb shortening with brachydactyly (Figure 1J,K,S,T,X,Y). An older sibling passed away in early infancy due to a congenital heart defect with no genetic testing performed. During early childhood, he suffered from chronic rhinosinusitis and recurrent chest infections which became less frequent with age. He developed hyperechoic kidneys, proteinuria, and chronic renal disease as well as retinitis pigmentosa later in childhood.

Individual 3.II.1 was a from non-consanguineous parents. Very short ribs, shorter long bones, polydactyly, cleft lip and palate, a complex congenital heart defect (persistent left superior vena cava, atrioventricular septal defect, hypoplastic left heart, aortic atresia), lobulated anterior tongue, absent brain falx and absent corpus callosum, cloacal malformation, cystic dysplastic kidneys (Figure 1L,U,V,Z) and pathognomonic radiological pelvis yielded a clinical diagnosis of ATD or SRPS.

Family 4 of unknown consanguinity whose first-born child was prenatally diagnosed with ATD or SRPS due to polydactyly, short ribs, narrow thorax, and short long bones. He was born with a congenital heart defect (double outlet right ventricle). Postnatally, the diagnosis was confirmed by the classic ATD/SRPS pelvis appearance in x-ray. The child suffered from severe respiratory distress in the neonatal period requiring mechanical ventilation and subsequently passed away due to cardiorespiratory failure.

### Effects of the *IFT74* exon 2 deletion on ciliary ultrastructure

Family 1 clinical symptoms suggestive of PCD associated with *IFT74* exon 2 deletions prompted us to perform nasal brush biopsies (Figure 2). In fresh brushings, immunofluorescence analysis using acetylated tubulin as ciliary axonemal marker showed reduced numbers of very short motile cilia in all affected individuals as compared to controls. The shortened and sparse cilia phenotype persisted after six weeks of culture, excluding secondary effects due to infections (Figure 2A). TEM of the brushings from Family 1 confirmed the phenotype of very sparse cilia (Figure 2Ca,Cb). In contrast to the 10 micron length typical of normal cilia, *IFT74* mutant cilia were 0.5-1 microns long and failed to extend beyond the microvilli layer (Figure 2Cb). Basal body amplification and docking appeared unaffected in all three individuals with *IFT74* exon 2 deletion and no obvious defects affecting distal appendages or ciliary rootlets were visible (Figure 2Cc-Cf). TEM cross sections showed that the *IFT74* exon 2 deletion yielded a variety of defects including loss of central pair or outer doublet microtubules, microtubule translocations and loss of microtubule integrity (Figure 2Ci) as compared to control cilia (Figure 2Cj).

To understand how the loss of exon 2 affects IFT74 structure, we performed western blot analysis of fresh nasal brushing samples from family 1. Brushings from control patients showed a band of ∼69kD that likely represents full length IFT74. Affected individuals 1.II.1 and 1.II.2 lacked the 69kD band but had two or three smaller bands in the 50-60kD range. Individual 1.II.1 also showed a larger band not seen in the control or her sibling 1.II.2 (Figure 2B).

### Generation of mouse Ift74 alleles

To characterize the function of Ift74 in mouse, we obtained the *Ift74*^*Tm1a*^ allele from Jackson lab (Figure 3, 4, 5). This allele contains a promotor-less beta-galactosidase gene trap cassette and a neomycin selectable marker inserted into intron two along with Cre recombination sites flanking exon three (Figure 3A). The beta-galactosidase-neomycin insert in intron 2 is flanked by Frt recombination sites and can be removed with FlpE recombinase to create a floxed allele (*Ift74*^*Tm1c*^). Exon 3 can be excised with Cre recombinase. If this is done before the FlpE recombination, the beta-galactosidase gene trap cassette remains (*Ift74*^*Tm1b*^) (Figure 3, 6). If Cre combination occurs after FlpE recombination, then exon 3 will be removed leaving only Frt and a Cre recombinase sites in the intron between exons 2 and 4 (*Ift74*^*Tm1d*^) (Figure 3, 7). The Tm1a allele is expected to be a null or strong hypomorphic allele due to the gene trap cassette terminating transcription after exon 1. The Tm1b allele retains the gene trap cassette and lacks exon 3 suggesting that it should be at least as strong as the Tm1a allele. The Tm1c (flox) allele is expected to be normal and the Tm1d allele is designed to produce an out of frame splice of exon 2 to exon 4 and should be a null allele.

**Figure 3:**
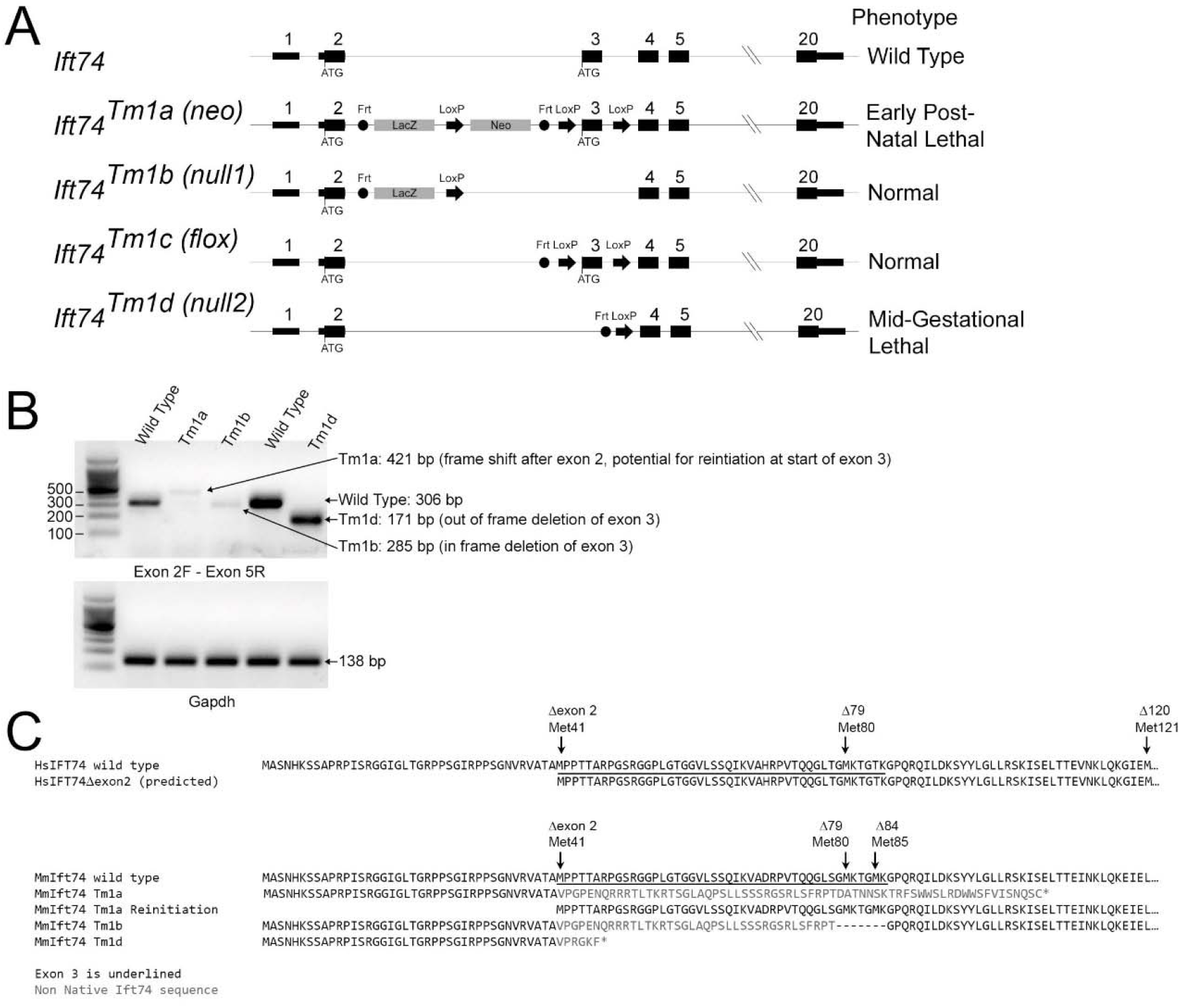
Mouse Alleles. **A**. Diagram of the alleles used in this study. The normal start codon in exon 2 and the potential alternative start codon at the start of exon 3 are shown. FRT and LoxP recombination sites are represented by circles and arrows respectively. Exons and introns are not drawn to scale and the introns do not reflect changes that occur during recombination. **B**. PCR amplification of cDNA generated from wild type, *Ift74*^*Tm1a*^, *Ift74*^*Tm1b*^ and *Ift74*^*Tm1d*^ variants using primers that span from exon 2 to exon 5. Note that alternative splicing produced small amounts of mRNA with the gene trap insertion removed from the *Ift74*^*Tm1a*^and *Ift74*^*Tm1b*^ alleles. The expected product was produced by the *Ift74*^*Tm1d*^ allele. Gapdh is included as a control for cDNA quality. Sequences of the products are included in Supplemental Data. **C**. Predicted protein sequence of the alleles.

**Figure 4:**
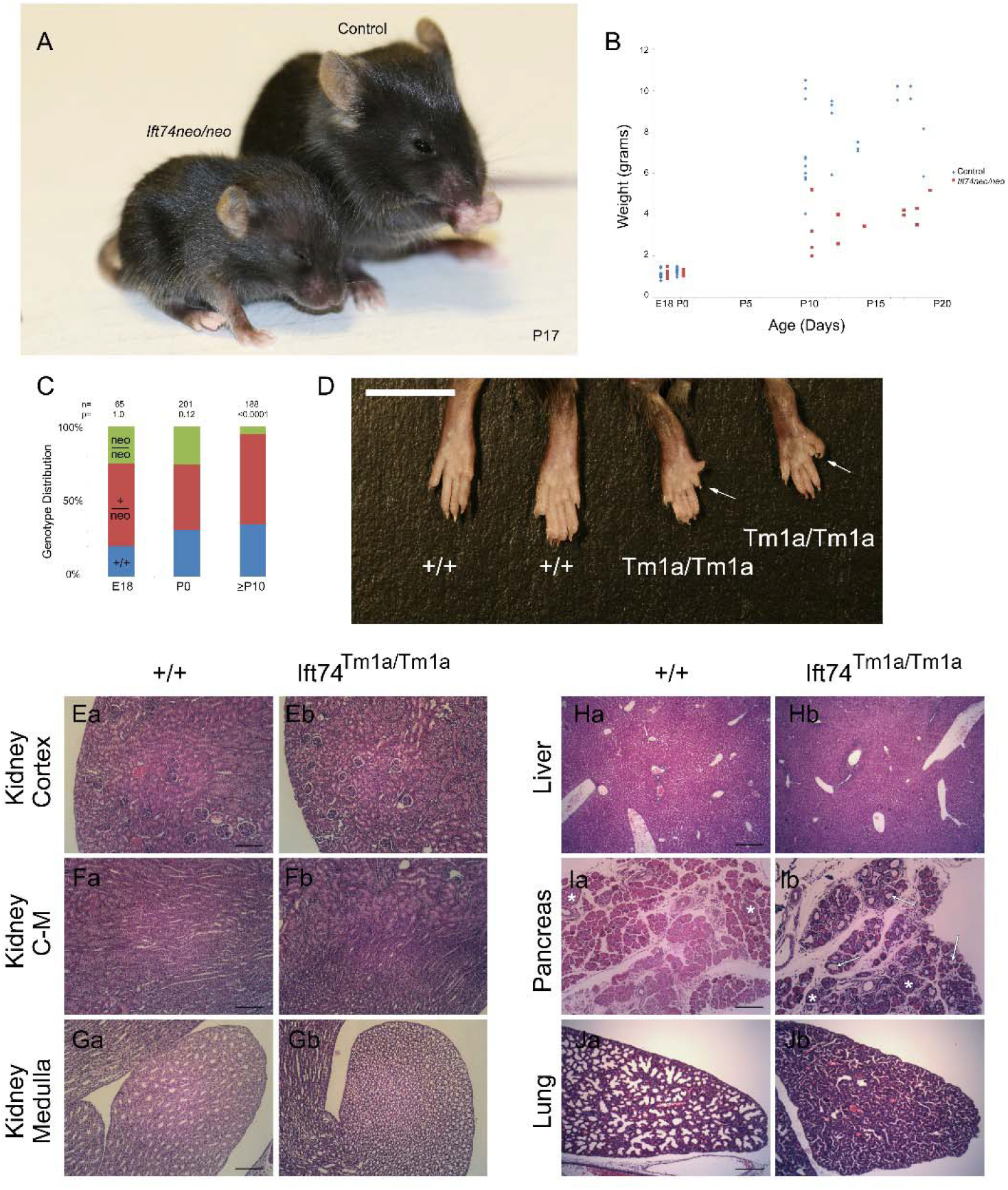
Ift74^Tm1a^ Mouse. **A**. Images of control and mutant mice at postnatal day 17. Note the smaller size and hydrocephaly. **B**. Weight of mice with respect to age. Each point represents a mouse at a particular age. Differences are significant with respect to genotype (P<0.0001, F test). **C**. Genotype distribution at the day prior to birth (E18), the day of birth (P0) and at genotyping age (P10 to P21). Number of animals is listed along with the P value determined by Chi Square analysis is listed at the top. **D**. Images of hindlimbs of controls and mutant animals. Note the extra digits on the mutants. **E-G**. H&E Images of P15 kidney cortex (control Ea, mutant Eb), cortical-medullary boundary (control Fa, mutant Fb) and medulla at the tip of the papilla (control Ga, mutant Gb). Scale bars are 100 microns. **H**. H&E Images of P8 liver (control Ha, mutant Hb). Scale bar is 200 microns. **I**. H&E Images of P8 pancreas (control Ia, mutant Ib). Islets are marked with an *. Arrows mark examples of cysts. Scale bar is 100 microns. **J**. H&E Images of P0 lung (control Ja, mutant Jb). Scale bar is 200 microns.

**Figure 5:**
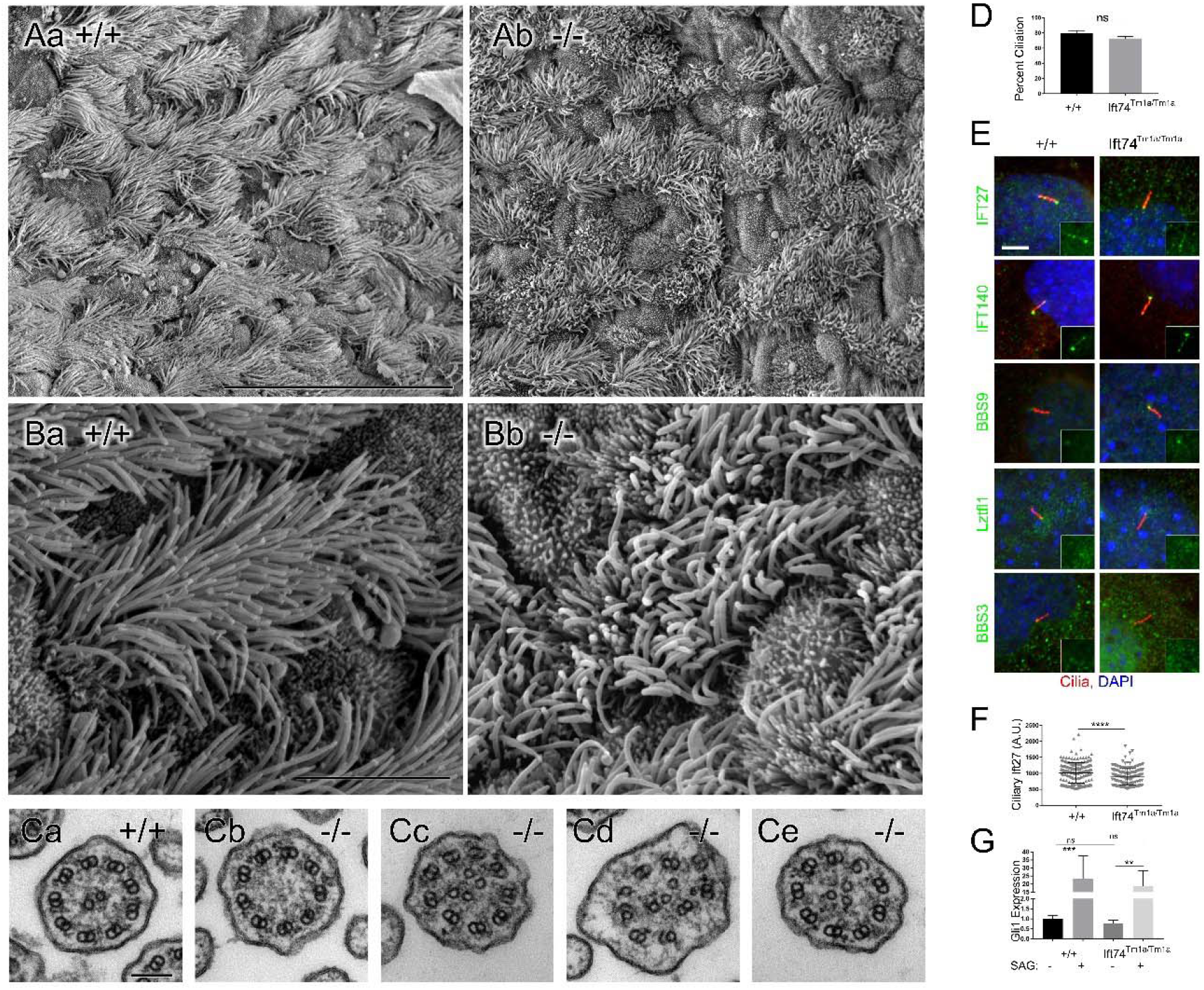
Ift74^Tm1a^ Cilia. **A, B**. Scanning EM of control (Aa, Ba) and *Ift74*^*Tm1a*^ mutant (Ab, Bb) trachea. Note the uneven length of the cilia in the mutant. Scale bar in A is 30 microns; in B it is 5 microns. N=4 mutant mice examined. **C**. Transmission EM of cilia from control (Ca) and *Ift74*^*Tm1a*^ mutant (Cb-Ce) trachea. Scale bar in G is 100 nanometers. Note the absence of central pair in Cb, the extra microtubules in Cc-Ce and the open B tubules in Cd and Ce. Quantification of ciliary defects is presented in Supplemental Material. **D**. Percent ciliation in serum starved MEFs. N=3 cell lines of each genotype, 100 cells counted per cell line. Difference is not significant (ns) by t-test. **E**. MEFs labeled with the IFT antibodies (green) as listed on the left side. Cilia are labeled with Arl13b (red). Quantification of the pattern is presented in Supplemental Material. **F**. Quantification of ciliary Ift27 levels (**** p<0.001 by t-test). **G**. Induction of *Gli1* by SAG. Differences between controls and the *Ift74*^*Tm1a*^ mutants are not significant (ns). Induction of Gli1 by SAG was significant in both genotypes (**p≤0.01, *** p≤0.001 by two-way ANOVA).

**Figure 6:**
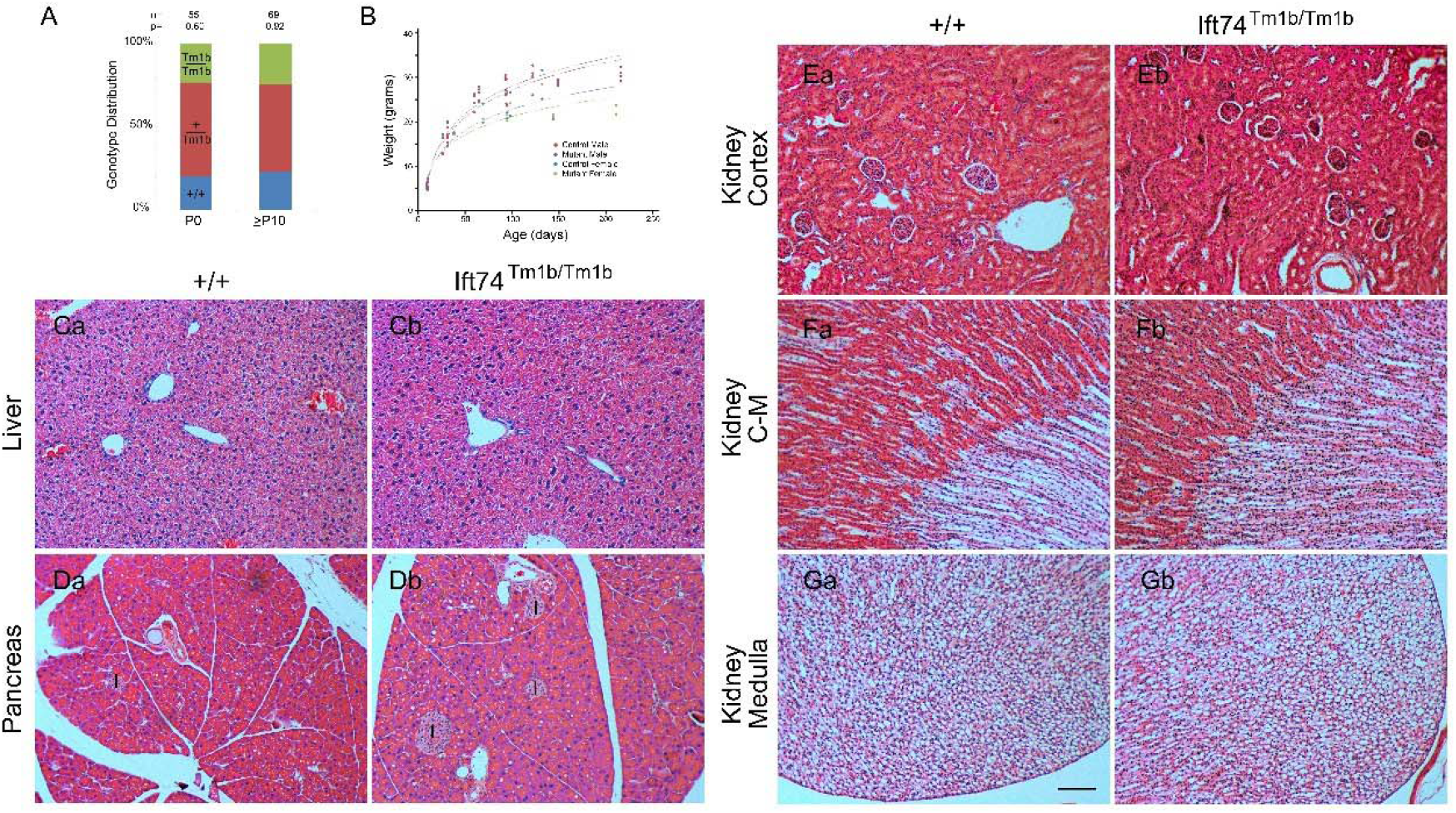
Ift74^Tm1b^ Mouse. **A**. Genotype distribution on the day of birth (P0) and at genotyping age (P10 to P21). Number of animals is listed along with the P value determined by Chi Square analysis is listed at the top. **B**. Weight of mice with respect to age. Each point represents a mouse at a particular age. The same mouse may be represented by multiple points. Genotype did not influence weight within each sex (p>0.05, F test). **C**. H&E Images of liver (control Ca, mutant Cb). **D**. H&E Images of pancreas (control Da, mutant Db). Islets are marked with I. **E-G**. H&E Images of kidney cortex (control Ea, mutant Eb), cortical-medullary boundary (control Fa, mutant Fb) and medulla at the tip of the papilla (control Ga, mutant Gb). Scale bar is 100 microns and applies to images in C-G.

**Figure 7:**
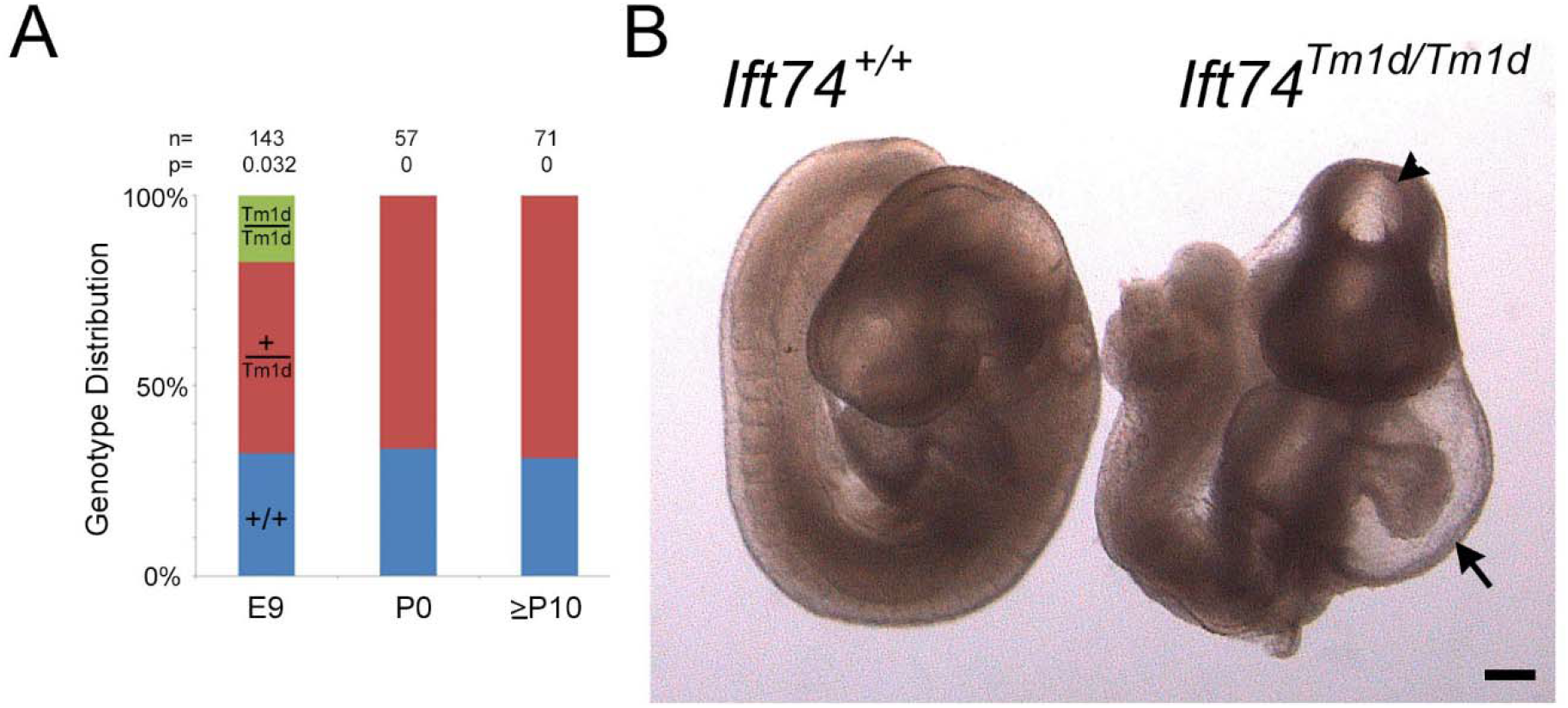
Ift74^Tm1d^ Mouse. **A**. Genotype distribution at embryonic day 9 (E9), the day of birth (P0) and at genotyping age (P10 to P21). Number of animals analyzed along with the P value determined by Chi Square analysis is listed at the top. **B**. Images of control and mutant mice at embryonic day 9. Note the malformed heart with pericardial effusion (arrow) and the abnormal head closure (arrowhead). Scale bar is 200 microns.

### *Ift74*^*Tm1a*^ causes postnatal lethality with polydactyly and hydrocephaly

Crosses between *Ift74*^*Tm1a*^ heterozygotes produced expected ratios of homozygous mutant animals on the day prior to birth (E18) and on the day of birth (P0) (Figure 4C). At E18 all mutants were alive and healthy appearing with polydactyly being the only obvious phenotype (Figure 4D). On P0, most of the mutants were dead by collection time. However, some survived and could be identified by the presence of polydactyly. Mutants usually lacked a milk spot indicating the failure to nurse. In typical large litters, none of the mutants survived past day 0 but with selective culling to reduce litter size to four, some mutants could survive past P0 with the oldest surviving to P20. The survivors were typically about half as big as littermates and developed hydrocephaly, necessitating euthanasia (Figure 4A,B).

Sixteen E18 and 23 P0 mutants were analyzed by microCT and necropsy (Table S2,S3[spreadsheets]). The prevalent phenotype observed by microCT was hindlimb polydactyly with most animals showing at least unilateral polydactyly. In addition, examples of duplex kidney and hydronephrosis were observed. Cardiac malformations were rare with only one example of a right aortic arch and a possible ventricular septal defect was observed.

Necropsy of P0 animals showed similar findings with a high penetrance of hindlimb polydactyly along with a four examples of forelimb polydactyly. Duplex kidney was observed in three animals along with one example of left lung isomerism and one example of malaligned sternal vertebrate. Significant disturbances of the left-right axis were not observed by either microCT or necropsy.

Histological analysis did not detect any evidence of cyst formation in the kidney or liver (Figure 4E-H). In contrast, every post P0 animal examined had an abnormally small pancreas. Histological analysis showed cyst formation within the exocrine ducts with fibrotic material surrounding the cystic ducts (Figure 4I). Lungs collected from two of four P0 animals showed reduced saccule area and abnormally thick walls suggesting hypoplasia (Figure 4J). The lung hypoplasia may be responsible for the inability of some animals to survive postnatally.

Hydrocephaly suggests motile cilia dysfunction, so we examined tracheal cilia by scanning and transmission EM (Figure 5). When examined by scanning EM, wild type tracheas showed uniform ciliation. In contrast, all mutants showed shorter and unequal cilia lengths as compared to littermate controls (Figure 5A). Transmission EM showed a variety of defects in cross sections of mutant cilia. These defects included absent central pair microtubules, extra central pair microtubules, displaced doublets, and failure of the B-tubule to fully connect to the A tubule (Figure 5C, S3).

MEFs derived from the *Ift74*^*Tm1a*^ mutant animals were able to ciliate as well as controls (Figure 5D) and staining with IFT markers, Ift140, Bbs9, Lztfl1 and Bbs3 did not show any differences from controls (Figure 5E, S3). The only detectable phenotype was a slight reduction in the amount of Ift27 localized to the base of the cilium (Figure 5E,F). Ift27 is thought to connect to the IFT particle through Ift74 and Ift81 and so this may reflect the reduction of Ift74 in the cells. This reduction does not appear to be highly significant as induction of Gli1 expression by SAG treatment was similar in mutants and controls (Figure 5G). Likewise, we did not observe any significant differences for CBY or CP110 localization in mutant MEFs compared to control MEFs (Figure S5).

### *Ift74*^*Tm1b*^ does not cause detectable phenotypes

Crosses between *Ift74*^*Tm1b*^ heterozygotes produced expected ratios of homozygous mutant animals on the day of birth (P0) and when genotyped at P10 or later (Figure 6A). Homozygous mutants were similar in weight to controls (Figure 6B), had no observable phenotypes and bred normally. Histological analysis detected no cysts or other pathology in the kidney, liver, or pancreas (Figure 6C-G). MEFs ciliated normally. These findings are similar to what was reported by the KOMP/EUCOMM phenotyping project (http://www.mousephenotype.org/data/experiments?geneAccession=MGI:1914944) where they reported abnormal blood glucose levels and behavioral differences but no structural birth defects or alterations in viability in this allele.

### *Ift74*^*Tm1d*^ caused mid-gestational lethality

Crosses between *Ift74*^*Tm1d*^ heterozygotes produced no homozygous mutant animals that survived to birth (Figure 7A). Other complex B null variants cause lethality starting at ∼E9 (34, 35). At E9, homozygous *Ift74*^*Tm1d*^ mutant embryos were slightly underrepresented (expected 36, obtained 26, P=.032). The mutants were all smaller than littermates, most failed to complete embryonic turning, had head closure defects and showed abnormal heart development with large pericardial effusions (Figure 7B). Most showed no evidence of heartbeat, and we were able to generate fibroblast cell lines from only one of seven embryos attempted while control littermates produced cell lines in all cases. The lack of heartbeat and failure to generate cell lines suggests that most embryos were already dead at the time of collection. Fibroblasts derived from the one mutant animal were completely unable to ciliate, but this could be rescued by transfection with Flag-tagged Ift74 (Figure 8A-C).

**Figure 8:**
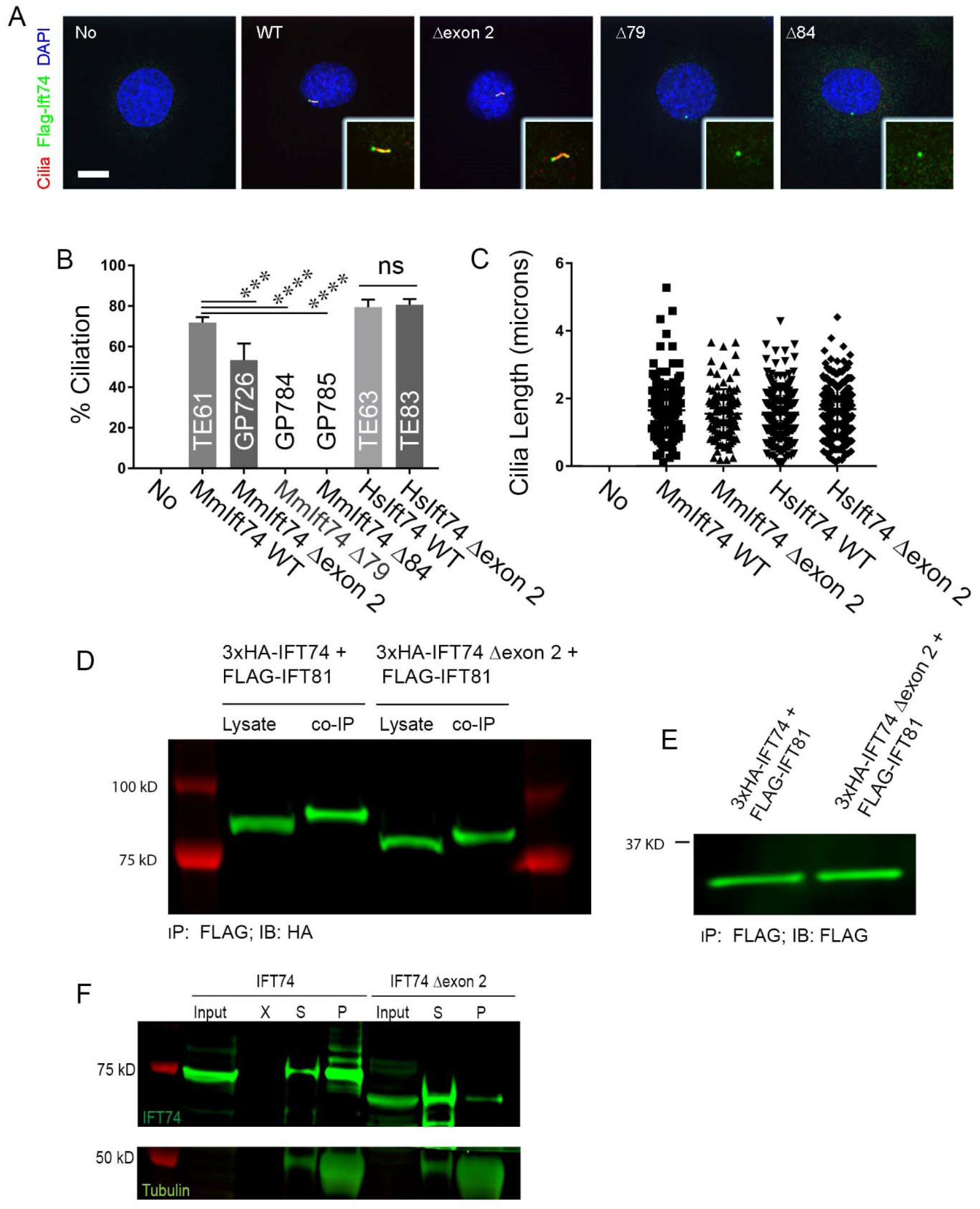
Ift74^Tm1d^ MEFs rescued with N-terminal deletion alleles. **A**. 20615.4T (*Ift74*^*Tm1d/Tm1d*^) MEFs derived from E9 mutant embryos were completely unable to ciliate (first column). Ciliation was rescued with wild type mouse *Ift74* (TE61), mouse *Ift74* lacking exon 2 with translation initiating at the second methionine (GP726), mouse *Ift74* initiating translation at methionine 3 (GP784), mouse *Ift74* initiating translation at methionine 4 (GP785), wild type human *Ift74* (TE63) or human *Ift74* lacking exon 2 (TE83). Mouse *Ift74* lacking exon 2 was less effective at rescuing ciliation (***p=0.004) but no difference was seen with human *Ift74* lacking exon 2. Mouse Ift74 lacking sequences upstream of the third and fourth methionines did not rescue (****p<0.0001) but did localize to the centrosome. N=100 or more cells counted from 3 independent experiments. Data was compared using one-way ANOVA. **B**. Cilia length in the same cells as described in A. No differences were seen in cilia length between any of the rescued lines that ciliated using one-way ANOVA. N=100 or more cilia measured per line. **C**. 20615.4T (*Ift74*^*Tm1d/Tm1d*^) MEFs unrescued (No) or rescued with TE61 expressing wild type mouse *Ift74* (WT), GP726 expressing *Ift74* initiating translation at the second methionine (Met2), GP784 expressing *Ift74* initiating translation at the third methionine (Met3) or GP785 expressing *Ift74* initiating translation at the fourth methionine (Met4). Cilia are labeled with Arl13b (red), Ift74-Flag (green) and DAPI (blue). Insets are 2X enlargements of the cilia/centrosome regions. Images are maximum projections of 3 image Z stacks taken at 0.2-micron intervals. Scale bar is 10 microns. **D**. Co-Immunoprecipitation of wildtype 3xHA-IFT74 and 3xHA-IFT74 lacking exon 2 with Flag-IFT81, showing similar amounts of IFT74 in lysates and co-IP samples. Co-IP was performed using anti Flag antibody and the blot stained with anti-HA antibody. **E**. IFT81 expression control for the co-immunoprecipitation shown in (D) showing similar expression levels in cells expression wildtype or mutant IFT74. **F**. IFT74-tubulin pulldown revealing decreased tubulin binding of Ift74 lacking exon 2 compared to wildtype IFT74. Tubulin was pelleted by ultracentrifugation and pellet and supernatant samples probed for presence of tubulin and IFT74. The majority of tubulin as well as wildtype IFT74 was found in the pellet after centrifugation, indicating IFT74-tubulin binding while IFT74 lacking exon 2 was detected predominantly in the supernatant indicating a failure to bind tubulin.

### Complex splicing alters the *Ift74* alleles

As described in previous sections, the phenotype of the Tm1b allele did not match expectations. This allele lacks exon 3 and carries a gene trap insertion that should strongly reduce transcription beyond exon 2. Thus, we expected a null or strong hypomorphic phenotype, but no abnormalities were observed. To understand the causes of this unexpected phenotype, we isolated mRNA from fibroblasts derived from each of the lines and analyzed the message structure around the insertion site by reverse transcriptase PCR (Figure 3B). Amplification of wild type cDNA with an exon 2 forward primer combined with an exon 5 reverse primer produced a 306 bp product with the expected sequence. Using the same primers on the Tm1a allele produced a small amount of a 421 bp product. Sequence of this product indicated that it contained exon 2 sequence and 115 base pairs of the gene trap vector fused to exon three (Supplemental Data). Translation of this message would go out of frame in the gene trap sequence (Figure 3C). However, the first codon of exon 3 is a methionine. This methionine is in the original reading frame potentially allowing for reinitiation of translation. The Tm1b allele produced a small amount of a 285 bp product due to an in-frame deletion of exon 3 and part of the gene trap cassette. The translated protein has the 45 residues from Met41 to Lys86 replaced with 38 residues derived from the targeting vector sequence. The Tm1d allele produced the expected 171 bp product where exon 2 was fused out frame to exon 4 (Supplemental Data).

### Exon 2 deletion impairs tubulin binding

To examine the importance of the Ift74 N-terminus to primary cilia formation, we transfected *Ift74* null (*Ift74*^*Tm1d/Tm1d*^) MEFs with expression constructs where translation would start at the normal methionine or at one of the three methionines in exon 3 (Figure 8A-C). The parental null MEFs are unable to ciliate. Rescue with Flag-tagged wild type mouse *Ift74* restored normal length cilia to ∼70% of cells (Figure 8B,C). Rescue starting at the second methionine was also fairly effective (∼50%) but rescue starting at the third or fourth methionine was ineffective with no ciliated cells being found. The latter two constructs were expressed and localized to a spot near the nucleus, which is likely the mother centriole (Figure 8A). Transfection with human IFT74 and IFT74^Δexon2^ was able to restore the ability to form normal length cilia to the null cells (Figure 8B,C). This indicates that IFT74 translated from the second methionine at the beginning of exon 3 is at least partially functional.

To determine if the deletion of exon 2 affected the ability of IFT74 to be incorporated into the IFT particle, we co expressed HA-tagged CBY (as a negative control) or IFT-B subunits, IFT81, IFT46 and IFT52, with Flag-tagged IFT74 or IFT74^Δexon2^ and immunoprecipitated with HA antibody. As expected, CBY did not precipitate any IFT74 or IFT74^Δexon2^ while IFT81, IFT46 and IFT52 similarly precipitated both IFT74 and IFT74^Δexon2^ (Figure 8D,E, S4A,B) indicating that the deletion of exon 2 does not greatly affect the ability of IFT74 to interact with other IFT-B proteins.

Structural studies indicate that the coiled-coil domain of IFT74 interacts extensively with IFT81 and together the IFT74/IFT81 N-termini form a tubulin binding site for transport of this major axonemal protein into cilia. The tubulin-binding site of IFT74 is thought to be coded by the N-terminal basic region (20), which is partly missing from IFT74^Δexon2^. Since IFT81 is thought to be the major tubulin binding site with IFT74 providing stability to the binding (20), we used an *in vitro* assay without IFT81 being present (Figure 8 F). To do this, we incubated *in vitro* translated full length IFT74 or IFT74^Δexon2^ with purified bovine tubulin. After centrifugation, the relative amounts in the supernatant and pellet were compared. In the absence of microtubules, both IFT74 and IFT74^Δexon2^ were found in the supernatant indicating that they are not aggregated. As expected, microtubules pelleted. When microtubules were added to translated IFT74 or translated IFT74^Δexon2^ and the solution centrifuged at 100,000 g, most of the full length IFT74 sedimented with the microtubules whereas IFT74^Δexon2^ was mostly found in the supernatant. A similar assay with lower g forces confirmed that IFT74^Δexon2^ did not sediment as well as full length and showed that IFT74^Δ79^ and IFT74^Δ120^ did not sediment with microtubules (Figure S4C).

## Discussion

IFT74 is a subunit of the IFT-B complex (36) and in *Chlamydomonas* it is required for ciliary assembly (21, 37). In mammals, it was initially called CMG1/IFT71 and shown to localize to cilia on endothelial cells (38). Human and mouse IFT74 genes are predicted to encode two main isoforms of 600 and 372 residues in humans and 600 and 388 residues in mouse. The isoforms share common N-terminal ends and in humans are identical for the first 351 amino acids and in mouse for 352 amino acids. The N-terminal ∼90 residues is rich in lysines and arginines and is thought to work with IFT81 to form a tubulin binding domain facilitating most tubulin transport during ciliary assembly (20, 21). The basic region is followed by a helical SMC domain, which facilitates interactions with multiple other IFT-B subunits. Most extensive interactions are seen with IFT81, which are distributed along the non-basic region of the protein, but structural analysis shows interactions with IFT70, IFT88, IFT46, IFT22, IFT25 and IFT27 indicating that IFT74 is central to IFT particle structure (39).

In this work, we identified four novel human *IFT74* disease-causing variants and characterized three mouse *Ift74* alleles. The mouse phenotypes ranged from a severe mid gestational lethal phenotype in the *Ift74*^*Tm1d*^ allele, a post-natal lethal phenotype in the *Ift74*^*Tm1a*^ allele, to no detectable phenotype in *Ift74*^*Tm1b*^ allele. In the *Ift74*^*Tm1d*^ allele, the mid gestational lethal phenotype occurred at embryonic day nine when the mice presented with severe cardiac malformations. This phenotype is similar to other null alleles of critical *IFT-B* genes like *IFT88* (35). The severe cardiac phenotype is likely to reflect the importance of cilia to heart development where it plays critical roles in setting up the left-right axis and in regulating Hedgehog signaling for development of the chambers and vasculature (13). In humans, equivalent heart development would occur in the first trimester, making it unlikely that strong alleles of critical ciliary assembly genes will be identified in viable or later-term lethal patients. The *Ift74*^*Tm1b*^ allele was predicted to be null, but due to unexpected splicing, it results in an in-frame replacement of the second coding exon with a similar number of codons derived from the targeting vector. The residues that are missing are from an unstructured domain that is thought to be part of the tubulin binding domain (20) but are clearly not critical to protein function. Conversely, the *Ift74*^*Tm1a*^ allele must affect protein function as these animals have significant phenotypes including growth restriction, hydrocephaly, polydactyly, and post-natal lethality. In mRNA isolated from these mice, the first coding exon is missing, which is likely compensated by translation initiation at a later methionine codon. Interestingly, the first triplet of the second coding exon specifies methionine. *In vitro* studies suggest that protein initiated at this codon is functional for primary ciliary assembly in fibroblasts (Figure 8). This is consistent with the mouse phenotype where motile cilia are affected but only subtle primary cilia phenotypes are observed.

Similar to what we observed in mouse, the human variants that we and others recovered in *IFT74*, result in a range of phenotypes. Most strikingly, we identified three individuals from two families whose IFT74 genes were missing the first coding exon (Families 1 and 2) in whom the first coding exon was deleted. The breakpoints identified by genome sequencing in 1.II.1 indicate that the deletion does not affect the antisense *IFT74* gene or other genes within the genome. Ensembl indicates a CFTC site in the deleted region. Effects of CTCF site deletions are difficult to assess. However, it seems unlikely that deletion of this site contributes to the motile cilia phenotype observed in the human patients as the motile cilia phenotype is reproduced in the mouse model lacking the CTCF genomic deletion. We predict this deletion will produce an N-terminally truncated protein identical to what is produced by the mouse *Ift74*^*Tm1a*^ allele. Post-natal growth restriction was observed in the mouse and the patients presented with short stature that became more pronounced with age. This phenotype not typically observed in ATD (25) where body length is often short at birth but catch up growth is normal except for cases with end-stage kidney disease. In addition to the short stature, the patients presented with recurrent respiratory infections. Light and electron microscopy revealed significant defects in the motile cilia of the respiratory tract. These defects persisted after respiratory cell culture suggesting that it is a primary defect and not secondary to infection. The *Ift74*^*Tm1a*^ mice developed hydrocephaly, which is a common phenotype of motile cilia defects in mice. Furthermore, scanning EM of the trachea of the affected mice showed shorter and fewer cilia and TEM showed a variety of axonemal defects.

Individual 1.II.1 showed proteinuria and kidney biopsy revealed mild tubular and glomerular dilations in early childhood but progressed to end-stage kidney disease requiring dialysis in adolescence. Individual 2.II.2 likewise suffers from reduced renal function and cystic dysplastic kidneys were observed in fetus 3.II.1. Cystic kidney disease, with associated liver and pancreas cysts and fibrosis, are common phenotypes in mice with cilia dysfunction. However, we did not observe kidney or liver phenotypes in the mouse although the pancreas was small and mildly cystic and fibrotic. *Ift74*^*Tm1a*^ mice typically presented with polydactyly. This is typically observed in about 10% of ATD patients but was not observed in any of the patients with an exon 2 deletion we studied. Retinal degeneration was observed in patient 2.II.2. This is common in mice with IFT-B variants (40) but we did not examine the retinas of the *Ift74*^*Tm1a*^ mice.

The individual in Family 3 was compound heterozygous for a splice donor variant c.974+4A>G and the intronic variant g.26982280delG (c.305+1664delG). Minigene splicing analysis suggests that the c.974+4A>G variant causes exon 12 to be skipped leading to a 13 amino acid deletion followed by frameshift and a premature stop codon. Variant g.26982280delG is unusual in that it is located deep in the intron of the major transcripts and would not be expected to be pathogenic. However, it is located at c.106-3delG in a rare transcript that uses alternative exons 5 and 6. Minigene analysis indicates that this variant shifts the balance to make more of the rare transcript and less of the major transcripts. The rare transcript produces a very short protein that is not likely to be functional for IFT-B particle assembly.

In Family 4, a newborn (4.II.1) with SRPS or lethal skeletal ATD phenotype, carried the variant c.789+2T>G. This variant, which is predicted to alter the exon 10 donor splice site, is likely to cause a 21 amino acid deletion after amino acid 263. Alternatively, it could result in the inclusion of the intron leading to an insertion of >1500 amino acids into the open reading frame, which seems unlikely. Regardless, the deletion or insertion would occur at a site where IFT74 has crosslinks to IFT81, IFT70 and the IFT25/IFT27 dimer making it likely that the variant would affect the ability of the protein to be incorporated into the IFT particle. The severe lethal ATD/SRPS phenotype caused by this variant suggests that it is a strong hypomorph.

Ten human cases carrying biallelic *IFT74* variants have been published to date: four BBS cases, four JBTS cases, and two cases with asthenozoospermia (isolated male infertility) (Table S1). Three of four BBS cases carry the same splice site allele, c.1685–1G>T, which is expected to affect splicing of intron 19. In one case, the variant was homozygous (41) while in the other two cases, the splice site variant was in trans to a heterozygous deletion of exons 14-19 (42) or a frameshift variant, c.371_372del (43). Disrupting splicing of intron 19 could potentially result in the production of protein consisting of 561 IFT74 residues followed by an extension of some number of non-IFT74 residues, although more disruptive outcomes such as message or protein instability are possible. The deletion of exons 14-19 would be expected to produce a protein of 351 normal residues with an unknown C-terminal end while c.371_372del is expected to cause a frameshift at Gln124. A fourth BBS patient (44) was heterozygous for c.535C>G, which would cause a Gln179Glu substitution and c.853G>T, which would cause termination at codon 285.

The same Gln179Glu substitution variant that was found in the BBS patient (c.535C>G) was identified in four JBTS cases. One family, with two affected individuals, also carries a c.92delT variant that is expected to terminate translation at codon 285. The second family carries a c.306-24A>G variant that is expected to delete Ser103 to Arg135. The third family had c.85C>T variant in trans that would be expected to cause termination after codon 29 (45). Additionally, two siblings with isolated male infertility were found to be homozygous for c.256G□>□A, which mutates Gly86 to Ser and changes the last nucleotide of exon 3. Changing the last nucleotide of exon 3 could potentially affect splicing. RT-PCR analysis showed that the variant did indeed cause imprecise splicing as a smear of amplification products was observed above and below the expected product. The two most abundant products were a normally spliced message with the Gly86Ser variant and one containing a 10-codon in-frame deletion between Leu77 and Gly86, but other splice variants were found (46). The Gly86Ser variant is within the unstructured basic domain and is not likely to have a serious effect on the protein. The 10 amino acid deletion could be pathogenic, but it is within a region deleted in our phenotype-less *Ift74*^*Tm1b*^ mouse, indicating that this is not critical sequence. The failure to splice properly could reduce the amount of functional protein below a level critical for sperm development, but not low enough to affect other cilia. Observing male infertility in human patients is consistent with results in mouse where targeted deletion of *Ift74* in the testes results in sperm tail assembly defects and male infertility (47).

Human patients with variants in IFT-B encoding genes have been previously described for the subunits IFT27 (BBS) (48, 49), IFT52 (SRPS) (50, 51), IFT54/TRAF3IP1 (Senior-Loken syndrome) (52), IFT56/TTC26 (biliary, renal, neurologic, and skeletal syndrome) (53), IFT57 (orofaciodigital syndrome) (54), IFT80 (SRPS) (55), IFT81 (SRPS) (56), and IFT172 (SRPS) (57). Like what we observed in our IFT74 cohort, the phenotypes in these patients ranged from BBS-like phenotypes to skeletal dysplasias with additional organ involvement. The phenotypes likely reflect the strength of the allele but also the particular subunit involved. For example, we now understand that some subunits like IFT74 are critical to IFT-B complex formation and so strongly deleterious alleles of these genes will affect ciliary assembly and give more severe phenotypes. Others like IFT25, IFT27, and IFT56 are not needed for ciliary assembly but play important roles in BBSome trafficking and Hedgehog signaling and are thus more likely to cause BBS like phenotypes (58-60).

Our studies combined with previously published studies of IFT74 patients present complicated but illuminating insight into the IFT74 structure function relationship. Loss of *IFT74* in mice leads to a severe phenotype with significant cardiac malformations. This is likely due to the failure of the IFT-B complex to assemble as IFT74 makes extensive contacts with other IFT-B proteins. Interestingly, we observed severe congenital heart defects in three of the five ATD/SRPS individuals reported here: double aortic arch in 2.II.2, persistent left superior vena cava, atrioventricular septal defect, hypoplastic left heart and aortic atresia in 3.II.1 and double outlet right ventricle and hypoplastic left ventricle in 4.II.1 No such heart defects have been previously reported in published BBS-, JBTS- or asthenozoospermia cases. The most severe human patients show SRPS/lethal ATD. These are often compound heterozygotes where one allele may be null due to a splicing defect early in the transcript and the other is a strong but not null allele. Splicing and other variants near the 3’end of the transcript result in BBS while a subtle variant near the N-terminus leads to isolated male infertility. Deletions near the N-terminus yield very different outcomes. In the mouse, replacement of the second coding exon with random residues produced no detectable phenotype indicating that it had little effect on the protein. In contrast removal of the first coding exon in either mouse or humans resulted a complex phenotype combining ATD-/SRPS-like chondrodysplasia with features of mucociliary clearance / PCD disorders. While all published BBS- and JBTS cases as well as the two SRPS cases carrying splice variants exhibited polydactyly, this was not the case for the three ATD individuals carrying exon 2 deletion. Our data suggests that this variant reduces tubulin binding but does not disrupt interaction with IFT81. The reduced tubulin binding likely causes axonemal microtubule extension deficits. This deficit is seriously detrimental to motile cilia but only mildly affects primary cilia. This is likely explained by higher demands for tubulin transport in motile cilia that are typically longer than primary cilia and may need more regeneration and repair due to mechanical stress not seen in primary cilia.

## Data Availability

All data produced in the present work are contained in the manuscript

## Abbreviations

ATD: Asphyxiating Thoracic Dystrophy
BBS: Bardet–Biedl syndrome
CED: Cranioectodermal Dysplasia
DAPI: 4′,6-diamidino-2-phenylindole
E: embryonic day
JBTS: Joubert syndrome
MEFs: Mouse embryonic fibroblasts
P: postnatal day
PCD: Primary Ciliary Dyskinesia
SRPS: Short-Rib Polydactyly Syndrome

## Acknowledgments

We thank Dr. Jaime Rivera for assistance photographing embryos and Dr. Joachim List, University Hospital Freiburg for providing a control for video microscopy. We would like to thank all participating patients and their families as well as referring medical doctors.

MS acknowledges funding from the European Research Council (ERC) (ERC starting grant No. 716344 to MS) and from the Deutsche Forschungsgemeinschaft (DFG, German Research Foundation)—Project-ID 431984000–SFB 1453) and Germany’s Excellence Strategy CIBSS—EXC-2189—project ID 390939984 under Germany’s Excellence Strategy. GJP funding was provided by National Institutes of Health grant R01 GM060992.

The authors have no competing interests.

