## Supplemental Material for "*IFT74* variants cause skeletal ciliopathy and motile cilia defects in mice and humans"

#### Supplemental Abbreviations

ATD, Asphyxiating Thoracic Dystrophy;
BBS, Bardet–Biedl syndrome;
DAPI, 4′,6-diamidino-2-phenylindole;
DMEM, Dulbecco's modified Eagle's medium;
E, embryonic day;
FBS, fetal bovine serum;
FCS, fetal calf serum;
JBTS, Joubert syndrome;
MEFs, Mouse embryonic fibroblasts;
P, postnatal day;
PCD, Primary Ciliary Dyskinesia;
SRPS, Short-Rib Polydactyly syndrome

#### Experimental Procedures

##### Human Subjects

This study involved individuals with the clinical diagnosis of as ATD or SRPS. Diagnostic criteria included short ribs/narrow ribcage detected prenatally or postnatally with or without polydactyly and radiological signs such as trident acetabulum with spurs in pelvis x-rays in the first year of life, handlebar clavicles or cone shaped epiphyses after the first year of life). DNA samples were collected after obtaining informed patient or parental consent. This work was approved by the Ethics committee of the Institute of Child Health, University College London, London UK (GOSH R&D number 11MM03, REC number 08/H0713/82) the ethics committee Arnhem-Nijmegen, The Netherland (ethical approval no 2006-048) and the ethics committee of Freiburg University, Freiburg, Germany (votum no 122/20).

##### Mouse Breeding

The $Ift74^{Tm1a}$ mice were obtained from the KOMP project at Jackson Laboratory. These mice contain a genetrap insertion and selectable marker but have all exons intact. This allele was converted to the Tm1b allele by deleting the floxed exon with Stra8-iCre (stock 017490, Jackson Laboratory, Bar Harbor, ME, USA)(Sadate-Ngatchou et al., 2008), and to the Tm1c flox allele by deleting the genetrap insertion and selectable marker from the germline with C57Bl/6J congenic FlpE (Farley et al., 2000). The flox Tm1c allele was converted to the Tm1d allele by deleting the floxed exon from Tm1c with Stra8-iCre. See figure 3 for diagrams. All lines were C57Bl/6J congenics maintained by recurrent mating to wild type C57Bl/6J purchased from Jackson Laboratory (stock number 000664).

Mouse work was carried out at the University of Massachusetts Medical School with IACUC approval.

#### Sequencing

**Exome Sequencing**

Exomic sequences from DNA samples were enriched with the SureSelect Human All Exon 50 V.6 Kit and 100-bp paired-end sequences were generated on a Hiseq PE150 (Illumina, San Diego, CA, USA). Read alignment and variant calling were performed with GATK using default parameters with the human genome assembly hg19 (GRCh37) as reference.

Following alignment and variant calling, serial variant filtering was performed as previously described (Rehman et al., 2019; Schmidts et al., 2013) for variants with a MAF equal or less than 0.5% in ExAc, 1000 genome project, esp6500 databases and gnomAD, coding variants or variants within 20 bp of exon-intron boundaries, and genes carrying bi-allelic variants with prioritization of homozygous variants in consanguineous pedigrees and genes with compound heterozygous variants in non-consanguineous pedigrees. Obligate loss of function variants such as canonical splice variants, frameshift and stop variants were prioritized over missense variants, however missense variants were not excluded from the analysis. BAM files were visually inspected for homozygous CNVs in all genes known to cause JATD or SRPS. CNV analysis was performed using ExomeDepth (Plagnol et al., 2012).

**Genome sequencing and break point PCR**

Genome sequencing from DNA sample of individual 1.II.1 was performed using the TruSeq DNA PCR-Free Kit (Illumina, San Diego, CA, USA) for library preparation which was sequenced as 2x150bp reads on a HiSeq4000 instrument (Illumina, San Diego, CA, USA). Read alignment and variant calling were performed using the GATK best practice pipeline with the human genome assembly hg19 (GRCh37) as reference. Reads were visualized using the Integrative Genomics Viewer (IGV) to determine the breakpoints. Subsequently, breakpoint junctions were amplified with the primers F1: TCTGAAATGAGGGTCCAGCTA, R1: GGGTGTTTAAGAACTCTCCTCCT and F2: AGAAATGACAACTGGTAAAAACATT, R2: ATGACGAGTTAGTGGGTGCAG. Resultant fragments were separated by agarose gel electrophoresis and sequenced by Sanger dideoxy nucleotide sequencing.

**Sanger sequencing**

Genomic DNA was isolated by standard methods directly from blood samples by Qiagen kit (Qiagen, Hilden, Germany). Amplification of genomic DNA was performed in a volume of 50 µl containing 30 ng DNA, 50 pM of each primer, 2 mM dNTPs, and 1.0 U GoTaq DNA polymerase (#M3001, Promega, Madison, WI, USA). PCR amplifications were carried out by an initial denaturation step at 94°C for 3 min, and 33 cycles as follows: 94°C for 30 sec, 58-60°C for 30 sec, and 72°C for 70 sec, with a final extension at 72°C for 10 min. PCR products were verified by agarose gel electrophoresis, purified, and sequenced bi-directionally. Sequence data were evaluated using the CodonCode software. Primer sequences are available on request.

**Splicing Assay**

Minigene assays for the c.974+4 A>G and (IFT74-006 ENSB0000578614-g.26982286delG, E5) alleles were performed as previously described (Maria et al., 2016). In brief, amplified DNA fragments of 444 bp for c.974+4 A>G including *IFT74* exon 14 and 801 bp for (IFT74-006 ENSB0000578614-g.26982286-delg, E5) including *IFT74* exon 5 along with their flanking intronic sequences were cloned between *RHO* exon 3 and exon 5 in pCI-neo mammalian expression vector and the plasmid transfected into HEK293T cells. After 48 hours of incubation at 37 °C, total RNA was isolated by Qiagen kit (Qiagen, Hilden, Germany) and analyzed by reverse transcriptase PCR (iSCRIPT, BioRad, Hercules, CA, USA). *IFT74* Primer sequences are available on request. The amplified fragments were electrophoretically separated on agarose gel followed by purification and Sanger sequencing.

#### Microscopy (Schmidts Lab)

**Transmission electron microscopy**

Samples are fixed using 3% glutaraldehyde (EM-grade). After fixation, the samples were washed in distilled water, osmicated for in 1% OsO4 in water, and dehydrated in a series of ethanol steps (70%, 80%, 90%, 96%) via propylene oxide and embedded in LX-112 resin. After polymerization at 60°C, ultrathin sections of 60-70 nm were cut on a Reichert EM UC6 using a diamond knife and collected on copper grids and stained with uranyl acetate and lead citrate. Images of the sections were produced using a FEI Tecnai-12 electron microscope at 100 kVe (Fei, Eindhoven, The Netherlands), with a Veleta camera and Radius software (EMSIS, Münster, Germany).

**High-speed videomicroscopy**

High-speed videomicroscopy was performed as previously described (Paff et al. 2017) using a Leica DM IRB microscope coupled to a digital camera (Basler A602F-2) with frame grabber (Matrox meteor II/1394). Videos were taken at 200 frames per second.

**Immunofluorescence Analysis**

In brief, cells were grown on coverslips or slides prepared from nasal brushings (cells grown on coverslips were serum starved for 24 hrs) and washed with PBS before they were fixed in 4% PFA for 5 minutes, washed 3x with PBS, permeabilized with PBS containing 0.05% Triton-X100, washed 3x in PBS, blocked in 5% BSA for 1 hour at room temperature or overnight at 4°C and then incubated overnight at 4°C in primary antibody 1:200 in 5% BSA in PBS. After washing for 5 times, secondary antibody solution was added (1:5000 in 5% BSA in PBS) and cells incubated for 2 hrs at room temperature. Slides and coverslips were mounted in Vectashield + 4′,6-diamidino-2-phenylindole (DAPI) (Vector Labs, Peterborough, UK). Images were taken with a Apotome Axiovert 200 (Zeiss, Oberkochen, Germany) and processed with AxioVision 4.8 (Zeiss, Oberkochen, Germany) or a confocal microscope (Olympus, Tokyo, Japan) and processed with Fiji (Imagej) software.

**Antibodies**

IFT74, rabbit polyclonal (HPA020247, Atlas antibodies, Bromma, Sweden),

CP110, rabbit polyclonal (12780-1-AP, Proteintech, Manchester, UK),

CBY, mouse monoclonal (8-2 sc-101551, Santa Cruz Biotechnology, Dallas, TX, USA),

ARL13B, rabbit polyclonal (17711-1-AP, Proteintech, Rosemont, IL, USA),

Acetylated-α-tubulin, mouse monoclonal (T7451, Sigma, Roedermark, Germany).

Anti-mouse Alexa Fluor 488 and anti-rabbit Alexa Fluor 546 were used as secondary antibodies (Molecular Probes Invitrogen, Carlsbad, CA USA).

#### Microscopy (Pazour Lab)

**Hematoxylin and Eosin**

Paraffin sections for H&E staining were dewaxed with Safeclear (Fisher Scientific, Hampton, NH) and rehydrated with graded aqueous solutions of isopropanol. The sections were stained for 4 min with CAT Hematoxylin (Biocare Medical, Concord, CA, USA), rinsed in running tap water for 30 sec followed by three quick dips in saturated lithium carbonate and a rinse in distilled water. This was followed by 90% ethanol for 2 min, Edgar Degas Eosin (Biocare Medical, Concord, CA, USA) for 2 min and 3 quick rinses in 100% ethanol. The sections were cleared with Safeclear (two 5 min incubations) and were mounted with Permount (Fisher Scientific, Hampton, NH, USA).

**Immunofluorescence of Tissues**

Paraffin sections were dewaxed, rehydrated and subjected to antigen retrieval in an autoclave (250°F, 40 min) with 10 mM sodium citrate at pH 6. Sections were brought to ambient temperature and treated with blocking solution (4% non-immune goat serum, 0.1% Triton X-100, 0.05% SDS, and 0.1% fish skin gelatin [Sigma] in TBST [0.05% Tween-20 in Tris-buffered saline, pH 7.4]) for 30 minutes, subsequently washed with TBST and then exposed to primary antibodies overnight at 4°C. Next day the sections were washed with TBST, incubated with Alexa Fluor-conjugated secondary antibodies (Life Technologies, Grand Island, NY, USA) for 30 min at 22°C, and washed with TBST followed by a rinse with TBS. The antibodies were brought to their working dilutions with 0.1% fish skin gelatin in TBS. The sections were then dipped for 5 seconds in DAPI (1 µg/ml in TBS) and after rinsing with TBS were mounted with Prolong Gold (Life Technologies, Carlsbad, CA, USA). Confocal images were acquired with an inverted microscope (TE-2000E2; Nikon, Tokyo, Japan) equipped with a Solamere Technology modified spinning disk confocal scan head (CSU10; Yokogawa, Tokyo, Japan). Z stacks were acquired at 0.5- micron intervals and converted to single planes by maximum projection with MetaMorph software (MDS Analytical Technologies, Sunnyvale, CA, USA).

**Electron Microscopy**

Tracheas were fixed with 2.5% glutaraldehyde, 2% paraformaldehyde in 100 mM cacodylate pH 7.2 overnight before osmication with 1% osmium tetroxide for 1 hr at 22°C. For transmission EM, the bottom half of the trachea was dehydrated through the following series: 10%, 25% (plus 4% uranyl acetate), 30%, 50%, 70%, 85%, 95% and 100% ethanol and then infiltrated first with two changes of 100% propylene oxide and then with a 50%/50% propylene oxide/SPI-Pon 812 resin mixture for overnight incubation. The following day three changes of fresh

100% SPI-Pon 812 resin were done before the samples were polymerized at 68°C in plastic capsules. Thin sections of approximately 70 nm were placed on copper support grids and contrasted with lead citrate and uranyl acetate. Sections were examined using the FEI Tecnai (Thermo Fisher, Waltham, MA, USA) 12 BT with 80Kv accelerating voltage, and images were captured using a Gatan (Pleasanton, CA, USA) TEM CCD camera.

For scanning EM, the top half of the trachea was dehydrated through the following series: 10%, 30%, 50%, 70%, 85%, 95% and 100% ethanol and then critical point dried in liquid $CO_2$. The tracheas were cut in half longitudinally and mounted onto aluminum stubs with silver conductive paint and coated with carbon (1 nm) and sputter coated with gold/palladium (12 nm). Specimens were examined using an FEI (Hillsboro, OR, USA) Quanta 200 FEG MK II scanning electron microscope.

**Immunofluorescence of Cells**

Cells for immunofluorescence microscopy were grown on acid-washed glass coverslips. The cells were fixed for 10 min in -20º C methanol or for 15 min in 2% paraformaldehyde, 0.05 M Pipes, 0.025 M Hepes, 0.01 M EGTA, 0.01 M MgCl2 (pH 7.2) followed by a two-min extraction with 0.1% Triton X-100 in the same solution. For some antibodies, an antigen retrieval step of 0.05% SDS in PBS for 5 minutes included at this point. After two brief washes in TBST (0.01 M Tris, pH 7.5, 0.166 M NaCl, 0.05% Tween 20), the cells were blocked with 1% bovine serum albumin (BSA) in TBST for 1 hr and then incubated with the primary antibodies either overnight at 4°C or for 2 hr at room temperature. The cells were then washed 4 times with 1% BSA/TBST over ~30 min. The cells were then incubated with 1:2000 dilutions of Alexa fluor-conjugated secondary antibodies (Molecular Probes, Eugene, OR, USA) for 1 hr and washed 4 times with 1% BSA/TBST over ~30 min followed by a brief wash with TBST. The cells were then mounted with ProLong Antifade (Molecular Probes, Eugene, OR, USA) and visualized by fluorescence microscopy. Images were acquired by an Orca ER camera on a Zeiss Axiovert 200M microscope equipped with a Zeiss 100X plan-Apochromat 1.4 NA objective by Openlab (Lexington, MA, USA) and adjusted for contrast in Adobe Photoshop. If comparisons are to be made between images, the photos were taken with identical conditions and manipulated equally.

Quantification of fluorescence intensity was performed using an ImageJ macro-toolset (Schneider et al., 2012) (https://imagej.net/ij/). Briefly, sets of images are imported into the software and cilia are pre-processed (median filtering and background subtraction), then detected using the “Analyze particle function”. Each cilium is extracted using a 64x64 pixels bounding box. Deconvolution is performed on both the structural marker channel and the signal channel with the DeconvolutionLab plugin (Sage et al., 2017), using synthetic PSF generated by the Diffraction PSF 3D plugin (http://www.optinav.info/Diffraction-PSF-3D.htm).

**Antibodies**

Arl13b mouse monoclonal N295B/66 (Neuromab, University of California Davis, CA, USA),
Gamma tubulin mouse monoclonal GTU88 (Sigma, St. Louis, MO, USA),
Ift27 (Keady et al., 2012),
Ift140 (Jonassen et al., 2012),
Bbs9 polyclonal (Sigma, St. Louis, MO, USA, catalog number HPA021289),
Lztfl1 rabbit polyclonal (Proteintech, Rosemont, IL, USA),

Bbs3 rabbit polyclonal (Jin et al., 2010) (Gift of M. Nachury),

Anti-mouse and anti-rabbit Alexa Fluor (488, 594, 633) secondary antibodies (Molecular Probes Invitrogen, Carlsbad, CA USA).

#### Protein Analysis

**MEF Western Blot**

For western blots, MEFs were lysed directly into denaturing gel loading buffer (Tris-HCl 125mM pH6.8, Glycerol 20% v/v, SDS 4% v/v, β-Mercaptoethanol 10% v/v, bromophenol blue). Western blots were developed by chemiluminescence (Super Signal West Dura, Pierce Thermo, Waltham, MA,USA) and imaged using a LAS-3000 imaging system (Fujifilm, Tokyo, Japan).

**Nasal Ciliated Cell Western Blot.**

Nasal ciliated cell samples were obtained by gentle brushing deep inside the nasal cavities using a cervical brush (Frankmed, Germany). Cells were transferred from the brush to a 15 ml tube containing 5 ml of Dulbecco's modified Eagle's medium (DMEM) supplemented with 10% fetal calf serum (FCS), 1% pyruvate and 1% antibiotic mixture of penicillin and streptomycin and tubes centrifuged at 1200 rpm for 5 minutes to pellet the cells. Cell pellets were washed 3x with PBS and dry pellets snap frozen in liquid nitrogen.

Nasal epithelial cells were lysed in lysis buffer (50 mM Tris-HCl [pH 7.5], 150 mM NaCl, 1% NP-40) supplemented with complete protease inhibitor cocktail (Roche, Risch-Rotkreuz, Switzerland) and subjected to gel electrophoresis using a 4%–12% Bis-Tris SDS-PAGE gel (NuPAGE Novex, Thermo Fisher, Waltham, MA, USA) and blotted overnight. Blots were stained with anti-IFT74 antibody and fluorescence analyzed on a LI-COR Odyssey 2.1 (Thermo Fisher) infrared scanner.

**Co-Immunoprecipitations.**

Proteins tagged with 3xHA and 3xFLAG epitopes were generated from Gateway entry clones by LR Clonase reaction. All clones were confirmed by sequence analysis to match appropriate RefSeq genes. Overexpression of tagged versions of the proteins in HEK293T cells, whole-cell extracts and western blots were conducted as previously described (Loges et al., 2018). Plasmids expressing N-terminal 3x Flag-tagged IFT74 were co-transfected with plasmids expressing N-terminal 3x HA-tagged IFT81, IFT46, IFT52 and CBY in HEK293T cells. 24 hrs after transfection, cells were lysed on ice using lysis buffer (50 mM Tris-HCl [pH 7.5], 150 mM NaCl, 1% NP-40) supplemented with complete protease inhibitor cocktail (Roche). Lysates were incubated with anti-HA affinity matrix (Roche) for 2–3 hr at 4°C. After incubation, beads with bound protein complexes were washed in ice-cold lysis buffer. Subsequently, 4X NuPAGE sample buffer was added to the beads and heated for 10 min at 70°C. Beads were pelleted by centrifugation, and supernatant was analyzed on NuPAGE Novex 4%–12% Bis-Tris SDS-PAGE gels. After blotting overnight at 4°C, blots were stained with mouse anti-FLAG or mouse anti-HA. Fluorescence was analyzed on a LI-COR Odyssey 2.1 infrared scanner.

**Tubulin binding assay.**

The microtubule binding assay was performed according to the procedure provided by Cytoskeleton Inc. Briefly, 20ul of bovine microtubules (Cytoskeleton MT-001m Denver, CO, USA) and 150 ug of wildtype or mutant protein *in-vitro* translated using TnT® SP6 Quick Master Mix Coupled Transcription/Translation System (Promega, USA), according to the manufacturer`s instructions or recombinant wildtype or mutant protein purchased from Genscript (Piscataway, NJ, USA) (BacPower Guaranteed Bacterial Protein Expression, >90% purity) were incubated for 30min at RT. The samples were then layered on cushion buffer (Cytoskeleton, Denver, CO, USA) to optimize microtubule polymerization (Cytoskeleton, Denver, CO, USA) and centrifuged at 100,000 g for 40 minutes.

#### Cell culture

**MEFs**

Mutant MEFs were isolated from E13.5 mutant embryos and immortalized with the large T antigen from SV40 virus. MEFs and their derivatives were grown at 37°C in 5% $CO_2$ in Dulbecco's modified Eagle's medium (DMEM; Gibco Thermo Fisher) with 5% FBS and 1% Penicillin-Streptomycin (Gibco Thermo Fisher). All DNA constructs were transfected into the cells via lentiviral infection followed by drug selection to create stable cell lines.

**Lentiviral Packaging**

Lentiviral packaged pHAGE-derived plasmids (Wilson et al., 2008) were packaged by a third generation system comprising four distinct packaging vectors (Tat, Rev, Gag/Pol, VSV-g) using HEK 293T cells as the host. DNA (Backbone: 5μg; Tat: 0.5 μg; Rev: 0.5 μg; Gag/Pol: 0.5 μg; VSV-g: 1 μg) was delivered to the HEK cells using calcium phosphate precipitates. After 48 hours, supernatant was harvested, filtered through a 0.45 mm filter and precipitated with Lenti-X concentrator (Clontech). Precipitated viral particles were resuspended in growth medium and added to ~50% confluent cells. After 24 hrs, cells were selected with antibiotic.

**Gli1 Expression**

For SAG experiments, cells were plated at near confluent densities and serum starved (same culture medium described above but with 0.25% FBS) for 48 hours prior to treatment to allow ciliation. SAG (Calbiochem) was used at 400nM. Isolation of mRNA and quantitative mRNA analysis was performed as previously described (Jonassen et al., 2008) using these primers.

| Primer | Accession No. | Sequence | Tm (C) | Amplicon |
|---|---|---|---|---|
| MmGAPDHExon3for | NM_008084 | GCAATGCATCCTGCACCACCA | 61.1 | 138 bp |
| MmGAPDHExon4rev | | TTCCAGAGGGGCCATCCACA | 61.1 | |
| MmGli1Exon4for | NM_010296 | CCAGGGTTATGGAGCAGCCAGA | 61.2 | 135 bp |
| MmGli1Exon5rev | | CTGGCATCAGAAAGGGGCGAGA | 61.5 | |

**Nasal Cell Culture**

Nasal cell culture was performed as previously described (Paff et al., 2017; Willems and Jorissen, 2004). In brief, nasal ciliated cells were obtained by gentle brushing deep inside the nasal cavities using a cervical brush (Frankmed, Germany. Cells were transferred from the brush to a 15 ml Falcon tube containing 5 ml of Dulbecco's modified Eagle's medium (DMEM) supplemented with 10% fetal calf serum, 1% pyruvate and 1% antibiotic mixture of penicillin and streptomycin. Cells were pelleted by centrifugation and treated with pronase over night before placement in a 25cm dish for 1 hr to remove fibroblasts. The supernatant was then transferred into a collagen coated T25 flask and cultured for three weeks. Upon confluency, cultures were treated with collagenase and cell sheets transferred into uncoated T25 flasks placed on a rotary shaker for one week followed by two weeks stationary incubation. Ciliated spheroids were kept in culture until used for imaging.

#### Plasmids

Plasmids were assembled using Gibson assembly (NEB, Ipswich, MA, USA) and the inserts and junctions verified by sequence. SnapGene files are available on request.

**TE61** MmIft74 (MASNHK…ASRS*). Mouse cDNA was codon optimized and chemically synthesized (IDT, Skokie IL, USA). This was amplified and cloned in a pHAGE-derived vector (Wilson et al., 2008) with an N-terminal 3X Flag tag. This construct is ampicillin resistant in bacteria and blasticidin resistant in mammalian cells.

**GP726** MmIft74Δexon2 Met 2(MPPTT...ASRS*). The first coding exon (second exon of gene) from TE61 was deleted using a PCR approach with Gibson assembly. Initiation begins at the second methionine at the start of the second coding exon. The N-terminal Flag tag was maintained.

**GP784** MmIft74 Met3 (MKTGMK...ASRS*). Nucleotides upstream from the third methionine were deleted from TE61 was deleted using a PCR approach with Gibson assembly. The N-terminal Flag tag was maintained.

**GP785** MmIft74 Met4 (MKGPQR...ASRS*). Nucleotides upstream from the fourth methionine were deleted from TE61 was deleted using a PCR approach with Gibson assembly. The N-terminal Flag tag was maintained.

**TE63** HsIft74 (MASNHK…TSGN*). The open reading frame of Hs*Ift74* was amplified from clone RC212777 (OriGene, Rockville MD, USA) and cloned into a pHAGE-derived vector with an N-terminal 3X Flag tag. This construct is ampicillin resistant in bacteria and blasticidin resistant in mammalian cells.

**TE83** HsIft74Δexon2 (MPPGT...TSGN*). The open reading frame minus the first coding exon (second exon of gene) was amplified from TE63 and cloned into the same vector as TE63. The N-terminal Flag tag was maintained.

#### Supplemental Table1: Summary of known *IFT74* variants

| | | | | | | |
|---|---|---|---|---|---|---|
| **ATD/PCD/SRPS** | Authors | Bakey et al. Pazour, 2023 | Bakey et al. Pazour, 2023 | Bakey et al. Pazour, 2023 | Bakey et al. Pazour, 2023 | Bakey et al. Pazour, 2023 |
| | Identifier | 1.II.1 | 1.II.2 | 2.II.2 | 3.II.1 | 4.II.1 |
| | Reference | This work | This work | This work | This work | This work |
| | Disease | ATD / PCD | ATD / PCD | ATD | ATD / SRPS | ATD / SRPS |
| | IFT74 Variant 1 | Exon 2 deletion (3kb) | Exon 2 deletion (3kb) | Exon 2 deletion (3kb) | c.974+4A>G (splice donor, intron 12) | c.789+2T>G ( splice donor, intron 10) |
| | IFT74 Variant 2 | Exon 2 deletion (3kb) | Exon 2 deletion (3kb) | Exon 2 deletion (3kb) | c.305+1664delG (splice donor, intron 4) | c.789+2T>G ( splice donor, intron 10) |
| **JBTS** | Authors | Luo et al. Cao 2021 | Luo et al. Cao 2021 | Luo et al. Cao 2021 | Luo et al. Cao 2021 | |
| | Identifier | 78C1 (Family 1_II:1) | 78C2 (Family 1_II:2) | 103C (Family 2_II:1) | 117C (Family 3_II:1) | |
| | Reference | PMID: 33531668 | PMID: 33531668 | PMID: 33531668 | PMID: 33531668 | |
| | Disease | JBTS | JBTS | JBTS | JBTS | |
| | IFT74 Variant 1 | c.92delT (p.Leu31Hfs*25) | c.92delT (p.Leu31Hfs*25) | c.306-24A>G [#] (p.Ser103_Arg135 del) | c.85C>T (p.Arg29*) | |
| | IFT74 Variant 2 | c.535C>G (p.Q179E) | c.535C>G (p.Q179E) | c.535C>G (p.Q179E) | c.535C>G (p.Q179E) | |
| | | | | [#]causes skipping of exon 5 resulting in an in-frame 33-amino acid deletion from Ser103 to Arg135 | | |
| **BBS** | Authors | Lindstrand et al. Katsanis 2016 | Kleinendorstvan et al. Haelst 2020 | Mardy et al. Slavotinek 2021 | Zhongling et al. Xiaoru 2021 | |
| | Identifier | | | | | |
| | Reference | PMID: 27486776 | PMID: 32144365 | PMID: 33748949 | PMID: 34539760 | |
| | Disease | BBS | BBS | BBS | JBTS | |
| | IFT74 Variant 1 | Deletion of exon 14–19 (Exon 14 starts with Gly352; exon 19 ends with Glu561) | c.371_372del (p.Q124Rfs*9) | c.1685-1G>T [#] (splice defect, intron 19) | c.535C>G (p.Q179E) | |
| | IFT74 Variant 2 | c.1685-1G>T [#] (splice defect, intron 19) | c.1685-1G>T [#] (splice defect, intron 19) | c.1685-1G>T [#] (splice defect, intron 19) | c.853G>T (p.E285*) | |
| | | [#]Exon 20 starts at Phe562. This variant could delete the last 39 residues. | | | | |
| **Male Infertility** | Authors | Lorès et al. Touré 2021 | Lorès et al. Touré 2021 | | | |
| | Identifier | patient 1 | patient 2 | | | |
| | Reference | PMID: 33689014 | PMID: 33689014 | | | |
| | Disease | Spermatogenic failure | Spermatogenic failure | | | |
| | IFT74 Variant 1 | c.256G > A [#] (p.Gly86Ser) and (p.Leu77_Gly86del) | c.256G > A [#] (p.Gly86Ser) and (p.Leu77_Gly86del) | | | |
| | IFT74 Variant 2 | c.256G > A [#] (p.Gly86Ser) and (p.Leu77_Gly86del) | c.256G > A [#] (p.Gly86Ser) and (p.Leu77_Gly86del) | | | |
| | | [#]Causes a splicing defect that produces aberrant messages. A common variant deletes amino acids Leu77 through Gly86 | | | | |

#### Supplemental Table 2

Spreadsheet of Ift74$^{Tm1a}$ MicroCT phenotypes

#### Supplemental Table 3

Spreadsheet of Ift74$^{Tm1a}$ necrospsy phenotypes

#### Supplemental Data: Sequence of MmIft74 cDNA between exons 2 and 5.

| | | |
|---|---|---|
| Primers: | MmIft74_exon2_for | ctcggggtggaataggactaacagg |
| | MmIft74_exon5_rev | gaagcttattaatttcagttgtaagttcac |

>MmIft74$^{wild\ type}$
Ctcggggtggaataggactaacaggaaggcctccttctggaataagacctccatctggcaatgttcgagtggcaactgcaatgccaccaacaacagcaagaccaggtt
ctcgtggtggtcccttagggactggtggagttttgtcatctcaaatcaaagttgctgatcgtcctgtgacccaacaaggtttgagtggaatgaagactggcatgaaaggtcc
ccagaggcaaattttagacaaatcttactatcttggacttcttaggagcaaaataagtgaacttacaactgaaattaataagcttc

>MmIft74$^{Tm1a}$
ctcggggtggaataggactaacaggaaggcctccttctggaataagacctccatctggcaatgttcgagtggcaactgcaGTCCCAGGTCCCGAAAAC
CAAAGAAGAAGAACCCTAACAAAGAGGACAAGCGGCCTCGCACAGCCTTCACTGCTGAGCAGCTCCA
GAGGCTCAAGGCTGAGTTTCAGACCAACAGatgccaccaacaacagcaagaccaggttctcgtggtggtcccttagggactggtggagtttt
gtcatctcaaatcaaagttgctgatcgtcctgtgacccaacaaggtttgagtggaatgaagactggcatgaaaggtccccagaggcaaattttagacaaatcttactatcttg
gacttcttaggagcaaaataagtgaacttacaactgaaattaataagcttc

>MmIft74$^{Tm1b}$
ctcggggtggaataggactaacaggaaggcctccttctggaataagacctccatctggcaatgttcgagtggcaactgcagtcccaGGTCCCGAAAACCA
AAGAAGAAGAACCCTAACAAAGAGGACAAGCGGCCTCGCACAGCCTTCACTGCTGAGCAGCTCCAGA
GGCTCAAGGCTGAGTTTCAGACCAACaggtccccagaggcaaattttagacaaatcttactatcttggacttcttaggagcaaaataagtgaactta
caactgaaattaataagcttc

>MmIft74$^{Tm1d}$
ctcggggtggaataggactaacaggaaggcctccttctggaataagacctccatctggcaatgttcgagtggcaactgcagtccccagaggcaaattttagacaaatctta
ctatcttggacttcttaggagcaaaataagtgaacttacaactgaaattaataagcttc

#### Supplemental Data: Predicted Ift74 protein sequence in human and mouse alleles.

>HsIFT74 Wild Type
**M**ASNHKSSAPRPISRGGIGLTGRPPSGIRPPSGNVRVATAMPPTTARPGSRGGPLGTGGVLSSQIKVAHRPVT
QQGLTGMKTGTKGPQRQILDKSYYLGLLRSKISELTTEVN…
>HsIFT74 Predicted deletion resulting from loss of exon 2
MPPTTARPGSRGGPLGTGGVLSSQIKVAHRPVTQQGLTGMKTGTKGPQRQILDKSYYLGLLRSKISELTTEV
N…

>MmIft74 Wild type
MASNHKSSAPRPISRGGIGLTGRPPSGIRPPSGNVRVATAMPPTTARPGSRGGPLGTGGVLSSQIKVADRPVT
QQGLSGMKTGMKGPQRQILDKSYYLGLLRSKISELTTEIN…
>MmIft74 Tm1a
MASNHKSSAPRPISRGGIGLTGRPPSGIRPPSGNVRVATAVPGPENQRRRTLTKRTSGLAQPSLLSSSRGSRL
SFRPTDATNNSKTRFSWWSLRDWWSFVISNQSC*
>MmIft74 Tm1a Reinitiation
MPPTTARPGSRGGPLGTGGVLSSQIKVADRPVTQQGLSGMKTGMKGPQRQILDKSYYLGLLRSKISELTTEI
N…
>MmIft74 Tm1b

MASNHKSSAPRPISRGGIGLTGRPPSGIRPPSGNVRVATAVPGPENQRRRTLTKRTSGLAQPSLLSSSRGSRL
SFRPT-------GPQRQILDKSYYLGLLRSKISELTTEIN…

>MmIft74 Tm1d

MASNHKSSAPRPISRGGIGLTGRPPSGIRPPSGNVRVATAVPRGKF*
GPQRQILDKSYYLGLLRSKISELTTEIN…

Methionines are in red text

Exon 3 is underlined

Non-native Ift74 sequence

#### Supplemental Data: Genotyping Primers

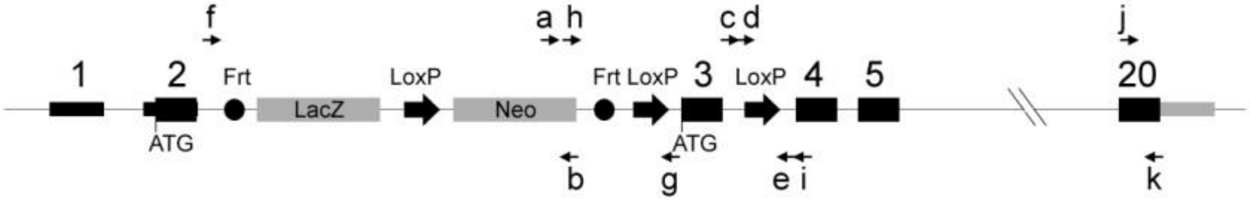


| | |
|---|---|
| a: LacZ Fwd | TACCGTTGATGTTGAAGTGGC |
| b: LacZ Rev | GACTGTAGCGGCTGATGTTG |
| c: IFT74_WT_Fwd.1 | GAATGCATGTGAAATACATTGTGAA |
| d: IFT74_Intron3_loxP_fwd.1 | CGCAATTAATGATAACTTCGTATAGC |
| e: IFT74_intron3_Rev | GAGAAAAGCAGTAATAGTTCTCATCTCC |
| f: IFT74_intron2_Frt_Fwd.2 | CTGAGTGAAAGTGGAGGC |
| g: IFT74_intron2_Frt2_Rev.3 | CAAGAAAGCTGGGTCTAGAT |
| h: IFT74_LacZ_SV40_Fwd2 | TAATAATAACCGGGCAGGGG |
| i: IFT74_Intron3_Rev.1 | GAGGGAAGCATAATGTCAGTC |
| j: 796_Ift74_F | CCTCACTTTATTTMAGAATGCAAGC |
| k:796_Ift74_R | GTTACAGAACAGAAACTGGTGCT |

a+b = LacZ band of approximately 300bp

c+e = Wild type band of approximately 400bp

d+e = Tm1a band of approximately 400bp

f+g = Tm1c band of approximately 200bp and wild type band of approximately 300bp

h+i = Tm1b band of approximately 700bp

e+f = Tm1e band of 539 bp and 1298 bp in wild type

j+k = 300 bp band that cuts in half with AciI when the *Ift74*$^{b2b796Clo}$ mutation is present

### Figure S1: Supplement to Support Figure 1.

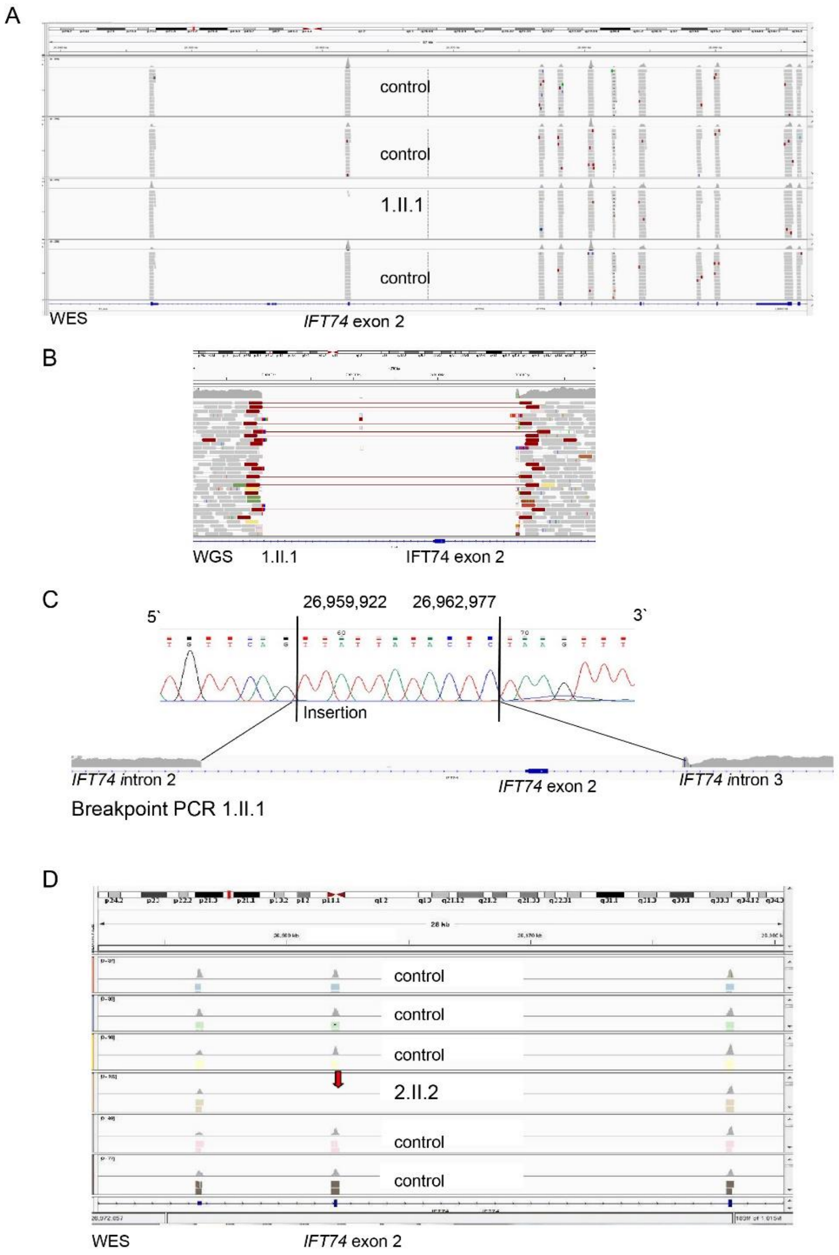


**A.** IGV screenshot of exome data from showing absence of sequencing reads in the patient 1.II.1 while this exon is well covered in controls. Other exons in the patient are well covered indicating that only exon 2 is deleted in the patient. **B.** IGV screenshot of *IFT74* exon 2 and surrounding intronic sequence from GS data in patient 1.II.1 showing the intronic breakpoints of the deletion. **C.** Confirmation of the intronic breakpoints by Sanger sequencing in Family 1 showing the deletion of *IFT74* exon 2 as well as parts of intron 1 and depicting a small insertion (TTATTATACTC). The intronic breakpoints are at 5`g.26,959,921 and 3`g.26,962,978. **D.** IGV screenshot of exome data from showing absence of sequencing reads in the patient 2.II.2 while this exon is well covered in controls. Other exons in the patient are well covered indicating that only exon 2 is deleted in the patient.

#### Figure S2: Supplement to Support Figure 1.

**A**

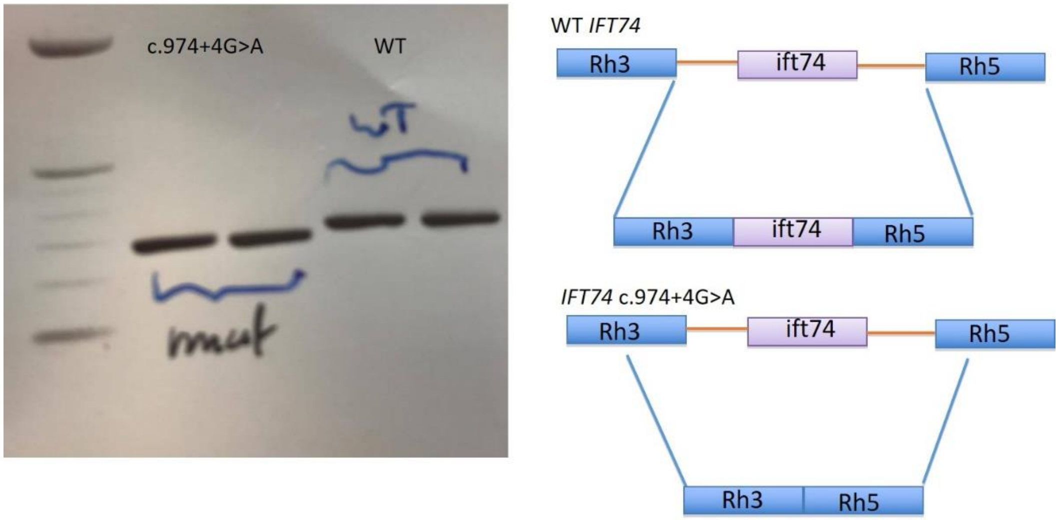


B

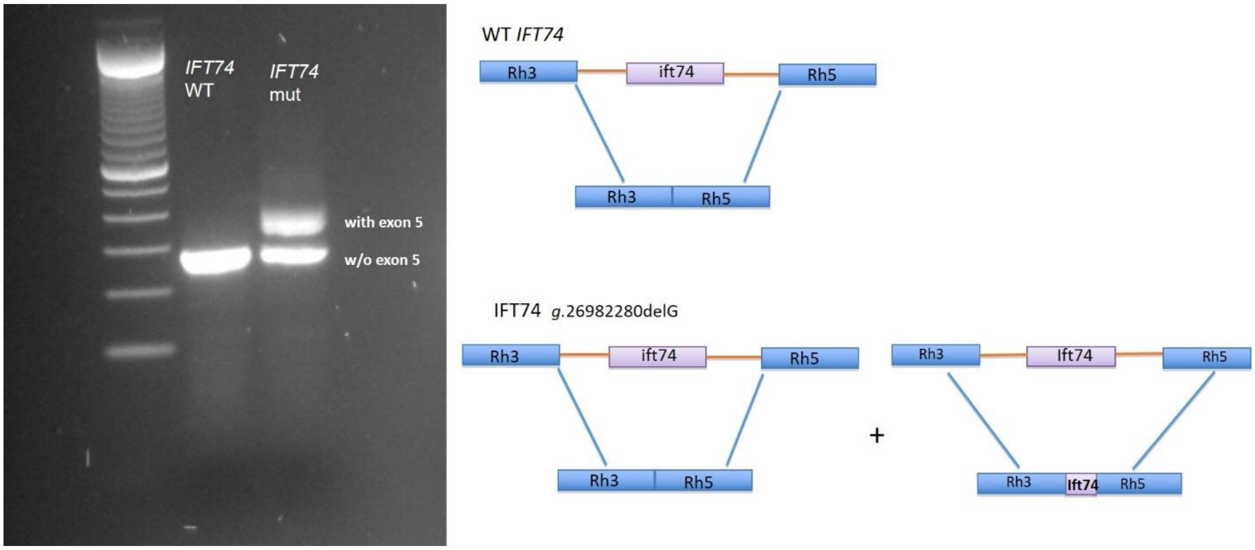


**A.** Minigene analysis of how splicing is affected by the c.974+4>A variant. Insertion of the wild type or the c.974+4>A version of *IFT74* exon 12 and surrounding intronic sequence between rhodopsin exons 3 and 5 resulted in product containing the rhodopsin exons spliced as expected to the *IFT74* exon while mRNA from the mutant lacked the *IFT74* exon. **B.** Minigene analysis of how splicing is affected by the g.26982280delG variant. The wild type or the g.26982280delG version of *IFT74* intron 4 including alternative exon 5 sequence was inserted between rhodopsin exons 3 and 5. Expression of this construct containing wild type sequence resulted in product containing rhodopsin exons 3 and 5 spliced with no IFT74 sequence included (w/o exon 5). The mutant form produced products with rhodopsin exons 3 and 5 spliced without exon 5 similar to wildtype and also produced a larger product with IFT74 alternative exon 5 (with exon 5) spliced between the two rhodopsin exons.

### Figure S3: Supplemental to Support Figure 5: Ift74$^{Tm1a}$ Cilia.

**Quantification of ciliary defects by TEM (see Figure 5C)**

Cilia from control and *Ift74$^{Tm1a}$* mutant trachea were imaged by TEM and ciliary defects counted

| Animal | Age | *Ift74$^{Tm1a}$* Genotype | Missing Central Pair | Super-numary MTs | Missing MTs | Displaced Outer Doublets | Unorganized Cilia | Norma l | Total | Percentage of defective cilia |
|---|---|---|---|---|---|---|---|---|---|---|
| 21297 | P19 | Mut | 1 | 4 | 2 | 1 | 2 | 110 | 120 | 8.3 |
| 21298 | P19 | Mut | 0 | 0 | 0 | 0 | 1 | 109 | 110 | 0.9 |
| 21300 | P19 | Het | 0 | 0 | 0 | 0 | 0 | 62 | 62 | 0 |
| 21301 | P19 | Het | 0 | 0 | 0 | 0 | 1 | 70 | 71 | 1.4 |
| 21798 | P15 | Mut | 0 | 14 | 12 | 3 | 9 | 189 | 227 | 16.7 |
| 21800 | P15 | WT | 0 | 2 | 0 | 0 | 2 | 274 | 278 | 1.4 |
| 21828 | P8 | Het | 0 | 0 | 0 | 0 | 0 | 147 | 147 | 0 |
| 21829 | P8 | Mut | 2 | 8 | 11 | 3 | 1 | 203 | 228 | 11.0 |

**Localization of IFT proteins to MEF cilia (see Figure 5E)**

Localization of IFT proteins in wild type and Ift74$^{Tm1a}$ MEF cilia. N=3 cell lines for each genotype, 25 cells examined from each cell line. P determined by student t-test.

IFT27 WT 100+/-0% spot at the base
MT 39+/-10% spot at the base
P=0.0005

IFT140 WT 100+/-0% spot at the base
MT 100+/-0% spot at the base
P=ns

BBS9 WT 57+/-10% spot at the base
MT 74+/-8% spot at the base
P=ns

Lztfl1 WT 100+/-0% no cilia label
MT 100+/-0% no cilia label
P=ns

BBS3 WT 100+/-0% no cilia label
MT 100+/-0% no cilia label
P=ns

#### Figure S4. Supplement to Support Figure 8

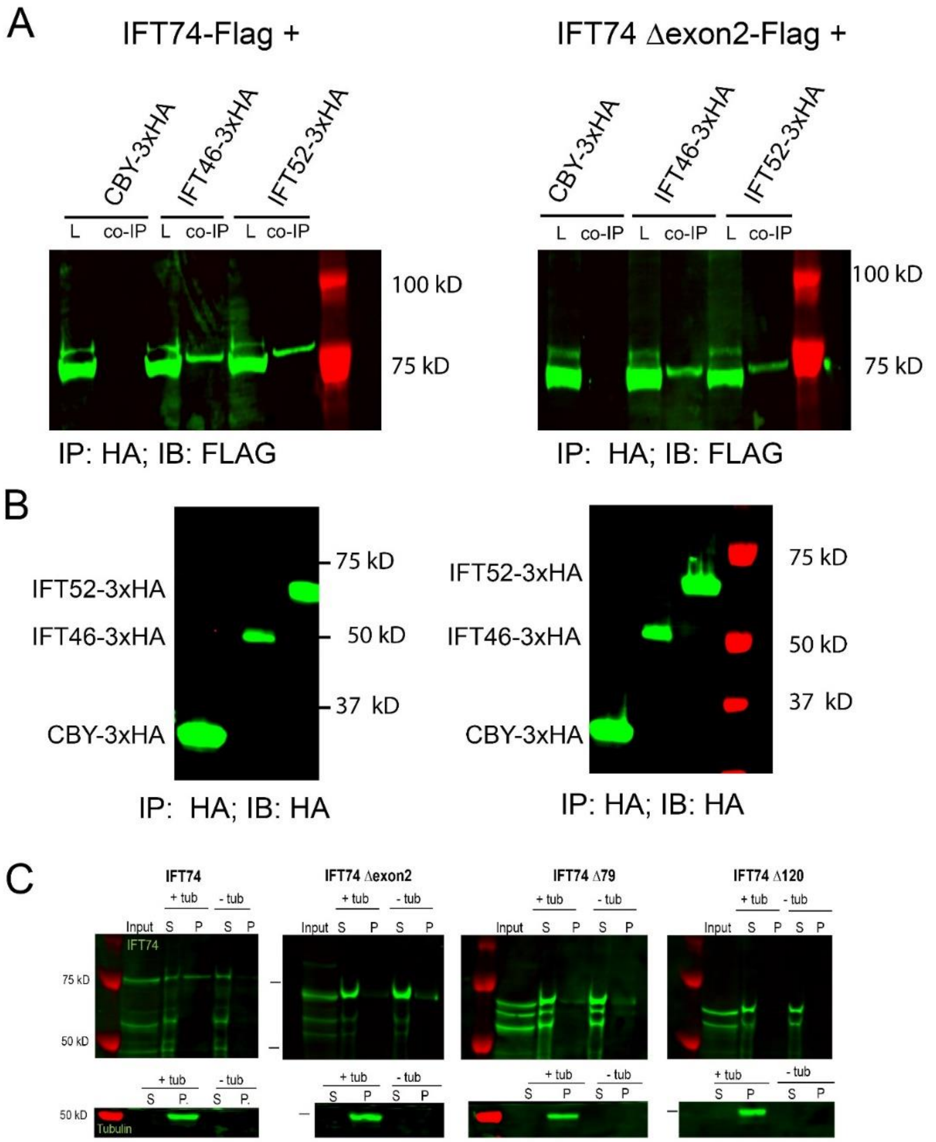


**A-B.** 3xHA-tagged human CBY, IFT46, and IFT52 were co-expressed in HEK293T cells with Flag-tagged IFT74 or IFT74$^{\Delta exon2}$. Extracts were precipitated with HA beads and the eluants probed for Flag (**A**) or HA (**B**). Note that IFT46 and IFT52 brought down similar amounts of IFT74 and IFT74$^{\Delta exon2}$ while no binding of wildtype or mutant IFT74 was observed with CBY. **C.** In vitro translated wildtype IFT74, IFT74$^{\Delta exon2}$, IFT74$^{\Delta 79}$, or IFT74$^{\Delta 120}$ was incubated with (+ tub) or without (- tub) microtubules and spun through a sucrose gradient. Note that microtubules pelleted (P) in all cases while wildtype or truncated IFT74 never pelleted without tubulin. Full length IFT74 was enriched in the pellet (P) compared to the soluble (S) fraction, indicating tubulin binding while truncated IFT74 was enriched in the supernatant, indicating impaired tubulin binding.

### Figure S5. Supplement to Support Figure 8

**A** **B**

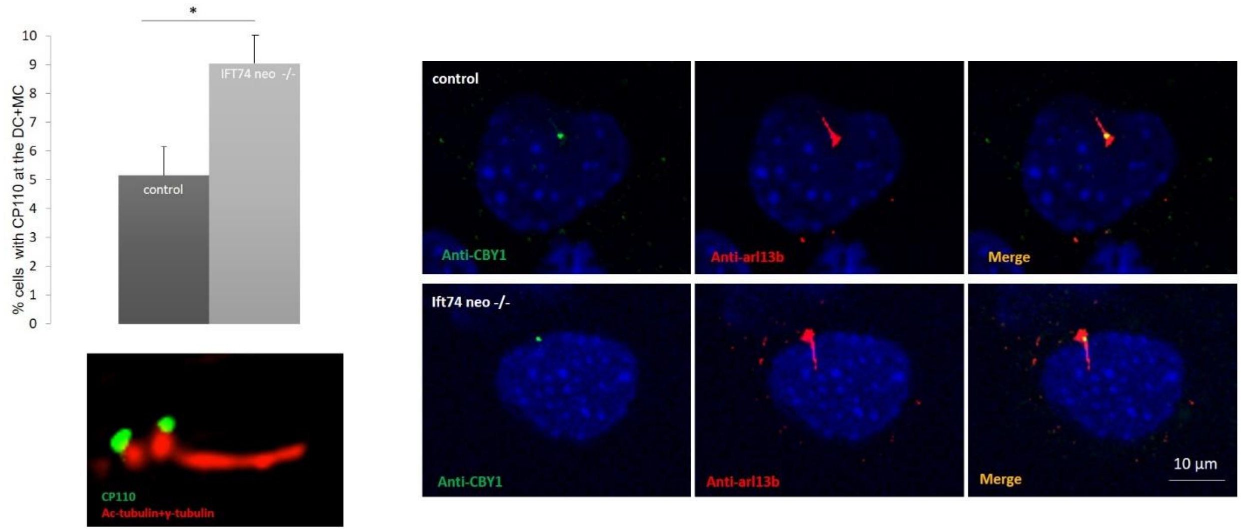


**A.** CP110 immunofluorescence analysis of cultured wildtype and IFT74 mutant MEFs reveals a slightly higher fraction of mutant cells with CP110 (green) present at both the mother and daughter centriole marked with gamma tubulin (red), ciliary axoneme marked with acetylated tubulin (red) (student t-test, $p< 0.05$), however over 90% of mutant cells showed no CP110 present at the mother centriole. **B.** Immunofluorescence analysis of CBY (green) revealed presence at the ciliary base in both wildtype and IFT74 mutant cultured MEFs. Ciliary axoneme marked with ARL13B (red).

### Supplemental Videos

Video 1: Live imaging of respiratory epithelia from 1.II.1 immediately after collection.

Video 2: Live imaging of respiratory epithelia from 1.II.2- immediately after collection.

Video 3: Live imaging of respiratory epithelia from 1.II.2 after explant was grown in culture for 6 weeks.

Video 4: Live imaging of respiratory epithelia from a control patient immediately after collection.
